# Genome-wide analyses identify 21 infertility loci and over 400 reproductive hormone loci across the allele frequency spectrum

**DOI:** 10.1101/2024.03.19.24304530

**Authors:** Samvida S. Venkatesh, Laura B. L. Wittemans, Duncan S. Palmer, Nikolas A. Baya, Teresa Ferreira, Barney Hill, Frederik Heymann Lassen, Melody J. Parker, Saskia Reibe, Ahmed Elhakeem, Karina Banasik, Mie T. Bruun, Christian Erikstrup, Bitten A. Jensen, Anders Juul, Christina Mikkelsen, Henriette S. Nielsen, Sisse R. Ostrowski, Ole B. Pedersen, Palle D. Rohde, Erik Sorensen, Henrik Ullum, David Westergaard, Asgeir Haraldsson, Hilma Holm, Ingileif Jonsdottir, Isleifur Olafsson, Thora Steingrimsdottir, Valgerdur Steinthorsdottir, Gudmar Thorleifsson, Jessica Figueredo, Minna K. Karjalainen, Anu Pasanen, Benjamin M. Jacobs, Nikki Hubers, Genes & Health Research Team, Estonian Biobank Research Team, Estonian Health Informatics Research Team, DBDS Genomic Consortium, FinnGen, Margaret Lippincott, Abigail Fraser, Deborah A. Lawlor, Nicholas J. Timpson, Mette Nyegaard, Kari Stefansson, Reedik Magi, Hannele Laivuori, David A. van Heel, Dorret I. Boomsma, Ravikumar Balasubramanian, Stephanie B. Seminara, Yee-Ming Chan, Triin Laisk, Cecilia M. Lindgren

## Abstract

Genome-wide association studies (GWASs) may help inform treatments for infertility, whose causes remain unknown in many cases. Here we present GWAS meta-analyses across six cohorts for male and female infertility in up to 41,200 cases and 687,005 controls. We identified 21 genetic risk loci for infertility (*P*≤5E-08), of which 12 have not been reported for any reproductive condition. We found positive genetic correlations between endometriosis and all-cause female infertility (*r*_g_=0.585, *P*=8.98E-14), and between polycystic ovary syndrome and anovulatory infertility (*r*_g_=0.403, *P*=2.16E-03). The evolutionary persistence of female infertility-risk alleles in *EBAG9* may be explained by recent directional selection. We additionally identified up to 269 genetic loci associated with follicle-stimulating hormone (FSH), luteinising hormone, oestradiol, and testosterone through sex-specific GWAS meta-analyses (N=6,095-246,862). While hormone-associated variants near *FSHB* and *ARL14EP* colocalised with signals for anovulatory infertility, we found no *r*_g_ between female infertility and reproductive hormones (*P*>0.05). Exome sequencing analyses in the UK Biobank (N=197,340) revealed that women carrying testosterone-lowering rare variants in *GPC2* were at higher risk of infertility (OR=2.63, *P*=1.25E-03). Taken together, our results suggest that while individual genes associated with hormone regulation may be relevant for fertility, there is limited genetic evidence for correlation between reproductive hormones and infertility at the population level. We provide the first comprehensive view of the genetic architecture of infertility across multiple diagnostic criteria in men and women, and characterise its relationship to other health conditions.

## Introduction

Infertility, defined as the inability to achieve pregnancy within 12 months of regular unprotected sexual intercourse, affects one in six couples across the globe^1^. A range of demographic, environmental, and genetic factors may drive infertility, including the age-related decline of sperm and oocyte quality and quantity^2,3^, infectious diseases^4–6^, and rare Mendelian disorders such as cystic fibrosis^7,8^. However, the exact cause remains undetermined in up to 28% of couples and 40% of women with infertility^9,10^. Given that current treatments such as *in vitro* fertilisation pose physical, emotional, and financial burdens on couples and healthcare systems^11–14^, a richer understanding of the biology and pathophysiology of infertility is urgently necessary.

Heritable women’s reproductive health diseases, particularly endometriosis^15^ and polycystic ovary syndrome (PCOS)^16^, are thought to be responsible for a considerable proportion of female infertility, with PCOS in particular accounting for up to 80% of cases of anovulatory infertility^17^. It is hypothesised that sex-hormone dysregulation^18,19^ and obesity^20^, which often accompany reproductive diseases, may be involved in the aetiology of infertility. Yet little is known about the genetic basis of reproductive hormones and infertility, which are not well-phenotyped in men or women in large studies^21,22^. Moreover, negative selection against infertility naturally limits the frequency of risk alleles in the population^23^. Genome-wide association studies (GWASs) have thus typically queried proxy measures of fertility such as childlessness^24,25^, which may partly arise from socio-economic and behavioural factors.

We aggregated data from a range of sources, including primary care and hospital electronic health records (EHRs) and self-report, across six cohorts with over 1 million participants, to perform the first reported GWAS meta-analyses for male infertility and five categories of female infertility. In addition, we report results from the largest sex-specific GWASs to date for five reproductive hormones. By aggregating this data with complementary rare variant genetic association testing from the UK Biobank, we catalogue the common and rare genetic contributions to infertility and reproductive hormone levels, quantify the extent of shared genetic architecture between these traits, and prioritise genes and cell types for further functional investigation of the hormonal and non-hormonal drivers of infertility.

## Results

### Genome-wide meta-analyses identify novel genetic loci for female and male infertility

We identified female infertility of all causes (F-ALL), anatomical causes (F-ANAT), anovulation (F-ANOV), unknown causes, i.e., idiopathic infertility as defined by exclusion of known causes of infertility (anatomical or anovulatory causes, PCOS, endometriosis, or uterine leiomyomas) (F-EXCL), or idiopathic infertility defined by inclusion of diagnostic codes for idiopathic infertility (F-INCL), as well as male infertility of all causes (M-ALL) in six cohorts, primarily of European ancestry (Figure 1 and Supp. Tables 1 and 2). The case-control ratio of all-cause female infertility ranged from 0.9% in the deCODE Genetics dataset^26^ to 11.7% in FinnGen^27^, whereas the case-control ratio of male infertility was between 0.3% (UKBB) and 8.2% (Danish Biobank) (Figure 1 and Supp. Table 2). Anatomical female infertility was the least common cause of infertility in three of six cohorts (prevalence in UKBB=0.01%, EstBB=2.0%, FinnGen=0.8%). Due to varying sample ascertainment, the case-control ratio does not necessarily reflect the population prevalence of infertility.

**Figure 1.**
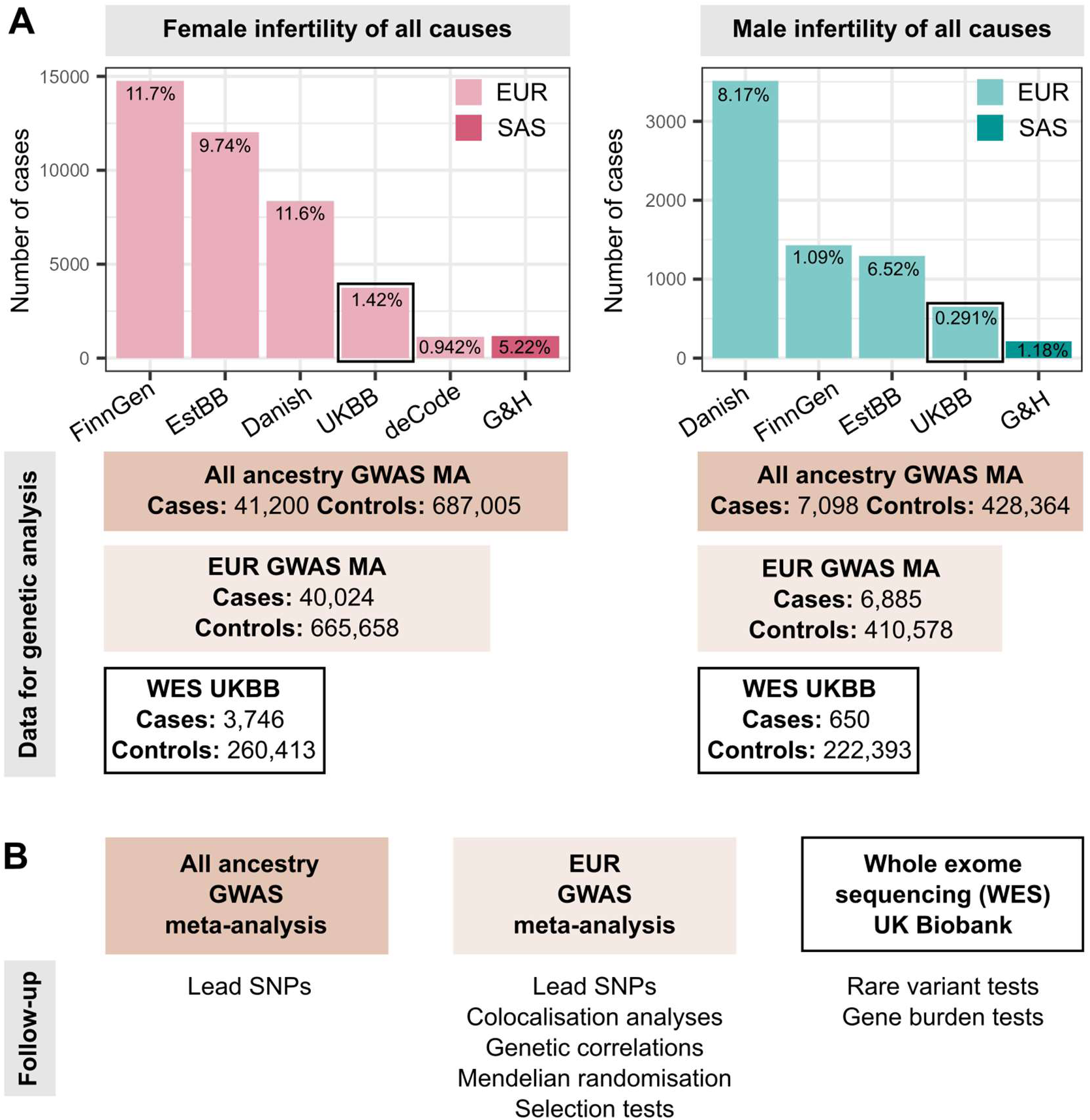
Overview of study cohorts and analyses presented for infertility genetic association studies. (A) Case numbers in each cohort contributing cases to genome-wide association study (GWAS) meta-analyses (MA) for female (left) and male (right) infertility. The prevalence of all-cause infertility in each cohort (%) is noted on the barplots. EUR=European ancestry, SAS=South Asian ancestry. EstBB=Estonian Biobank, Danish=Danish Blood Donor Study/Copenhagen Hospital Biobank, UKBB=UK Biobank, G&H=Genes and Health cohort. Total case and control counts for each type of genetic analysis: all ancestry GWAS meta-analysis (dark rectangles), EUR-only GWAS meta-analysis (light rectangles), and UK Biobank whole exome sequencing (WES) analyses (black outlined rectangles) are displayed. Male infertility in deCode, with <100 cases, was excluded from GWAS MA. Note the different Y-axis scales in each subplot. (B) Downstream analyses performed for each type of genetic analysis: lead variants were identified via distance-based pruning for all-ancestry and EUR-only GWAS meta-analyses; colocalisation, genetic correlation, and selection analyses were only performed for EUR meta-analyses due to the need for ancestry-matched linkage disequilibrium (LD) information; rare variant and gene burden tests were performed with WES data for the UK Biobank EUR-ancestry subset.

#### Novel genetic loci for infertility

We performed GWAS meta-analyses, testing up to 28.4 million genetic variants for associations with each of the above categories of infertility, in up to 41,200 cases/687,005 controls in women, and 7,098 cases/428,364 controls in men (Figure 1 and Supp. Table 2). We identified 19 unique genome-wide significant (GWS, *P*<5E-8) loci associated with at least one category of female infertility and two loci for male infertility (minor allele frequency (MAF) range 0.24%-46%, lead variants reported in at least two cohorts) (Figure 2, Table 1, and Supp. Figure 1). There was no evidence for heterogeneity in lead variant effects across cohorts (Supp. Text).

**Table 1.** Lead variants associated with infertility in GWAS meta-analyses. A1 is the effect allele. *lead variant is reported in only one cohort.

| RSID | chr:pos:A1:A2<br>(hg38) | Mapped<br>gene | All ancestries |  |  | EUR only |  |  |
| --- | --- | --- | --- | --- | --- | --- | --- | --- |
|  |  |  | Average<br>MAF | OR (95% CI) | P-value | Average<br>MAF | OR (95% CI) | P-value |
| Female infertility of all causes (F-ALL) |  |  |  |  |  |  |  |  |
| rs61768001 | chr1:22139327:T:C | WNT4 | 0.166 | 0.909 (0.891-0.927) | 2.25E-21 | 0.163 | 0.911 (0.893-0.93) | 1.24E-19 |
| rs10200851 | chr2:11581956:T:C | GREB1 | 0.458 | 0.951 (0.937-0.965) | 2.90E-11 | 0.456 | 0.951 (0.936-0.965) | 5.84E-11 |
| rs6938404 | chr6:151222906:T:C | AKAP12 | 0.453 | 0.958 (0.943-0.973) | 3.88E-08 | 0.453 | 0.958 (0.943-0.973) | 3.88E-08 |
| rs17803970 | chr6:152232583:A:T | SYNE1 | 0.0836 | 1.09 (1.06-1.12) | 1.91E-10 | 0.0836 | 1.10 (1.07-1.13) | 7.50E-11 |
| rs1964514 | chr8:109463457:C:G | EBAG9 | 0.0595 | 1.13 (1.09-1.16) | 3.01E-14 | 0.0597 | 1.13 (1.09-1.16) | 6.68E-14 |
| Anatomical female infertility (F-ANAT) |  |  |  |  |  |  |  |  |
| rs340879 | chr1:213983171:T:C | PROX1 | 0.418 | 0.906 (0.874-0.939) | 4.95E-08 | 0.418 | 0.902 (0.869-0.936) | 5.06E-08 |
| Anovulatory female infertility (F-ANOV) |  |  |  |  |  |  |  |  |
| rs72665317 | chr1:22040580:T:G | CDC42 | 0.190 | 0.875 (0.839-0.913) | 7.76E-10 | 0.18 | 0.886 (0.847-0.927) | 1.45E-07 |
| rs72827480 | chr2:120388925:T:C | INHBB | 0.401 | 0.905 (0.873-0.938) | 4.20E-08 | 0.401 | 0.905 (0.873-0.938) | 4.20E-08 |
| rs1852684 | chr2:145068818:T:G | ZEB2 | 0.367 | 1.12 (1.08-1.16) | 9.25E-10 | 0.35 | 1.12 (1.08-1.17) | 3.44E-10 |
| rs552953683 | chr8:102898586:T:C | AZIN1 | 0.0024 | 0.341 (0.234-0.498) | 2.54E-08 | 0.0024 | 0.341 (0.234-0.498) | 2.54E-08 |
| rs9696009 | chr9:123856954:A:G | DENND1A | 0.0777 | 1.21 (1.14-1.29) | 6.87E-10 | 0.0695 | 1.24 (1.16-1.32) | 2.40E-10 |
| rs9902027 | chr17:7537667:T:C | TNFSF12 | 0.255 | 0.895 (0.86-0.931) | 4.06E-08 | 0.255 | 0.895 (0.86-0.931) | 4.06E-08 |
| rs143459581 | chr22:28068862:T:C | PITPNB | 0.0419 | 1.30 (1.19-1.43) | 1.21E-08 | 0.0419 | 1.30 (1.19-1.43) | 1.21E-08 |
| rs17879961 | chr22:28725099:A:G | CHEK2 | 0.0389 | 0.739 (0.673-0.811) | 1.55E-10 | 0.0389 | 0.739 (0.673-0.811) | 1.55E-10 |
| Idiopathic female infertility, exclusion definition (F-EXCL) |  |  |  |  |  |  |  |  |
| rs61768001 | chr1:22139327:T:C | WNT4 | 0.165 | 0.923 (0.902-0.946) | 7.49E-11 | 0.162 | 0.928 (0.906-0.951) | 2.48E-09 |
| rs111597692 | chr8:109039973:T:C | TRHR | 0.0323 | 1.16 (1.10-1.22) | 1.51E-08 | 0.0323 | 1.16 (1.1-1.22) | 1.51E-08 |
| rs17378154 | chr8:109568721:A:G | EBAG9 | 0.059 | 1.13 (1.09-1.17) | 1.64E-10 | 0.0593 | 1.13 (1.09-1.17) | 3.36E-10 |
| Idiopathic female infertility, inclusion definition (F-INCL) |  |  |  |  |  |  |  |  |
| rs61768001 | chr1:22139327:T:C | WNT4 | 0.170 | 0.87 (0.839-0.903) | 6.87E-14 | 0.165 | 0.872 (0.840-0.905) | 8.96E-13 |
| rs11692588 | chr2:11544358:A:G | GREB1 | 0.358 | 0.919 (0.892-0.947) | 2.98E-08 | 0.358 | 0.919 (0.892-0.947) | 2.98E-08 |
| rs190290095 | chr4:39786858:A:G | UBE2K | 0.0022 | 0.227 (0.137-0.375) | 7.60E-09 | 0.0022 | 0.227 (0.137-0.375) | 7.60E-09 |
| rs851982 | chr6:151703850:T:C | ESR1 | 0.428 | 0.921 (0.895-0.947) | 7.60E-09 | 0.437 | 0.922 (0.896-0.949) | 2.86E-08 |
| rs17378154 | chr8:109568721:A:G | EBAG9 | 0.0565 | 1.18 (1.11-1.25) | 2.47E-08 | 0.0569 | 1.18 (1.11-1.25) | 4.97E-08 |
| rs74156208 | chr10:61509370:A:G | TMEM26 | 0.184 | 1.10 (1.06-1.14) | 4.96E-08 | 0.187 | 1.10 (1.07-1.15) | 5.44E-08 |
| Male infertility of all causes (M-ALL) |  |  |  |  |  |  |  |  |
| rs1228269928* | chr2:132923776:A:T | NCKAP5 | 0.0006 | 0.0995 (0.0441-0.224) | 2.72E-08 | 0.0006 | 0.0995 (0.0441-0.224) | 2.72E-08 |
| rs150639836 | chr10:53879806:T:C | PCDH15 | 0.0109 | 0.505 (0.402-0.636) | 5.72E-09 | 0.0109 | 0.505 (0.402-0.636) | 5.72E-09 |
| rs139862664 | chr10:116879589:C:G | ENO4 | 0.0072 | 0.388 (0.277-0.543) | 3.29E-08 | 0.0072 | 0.388 (0.277-0.543) | 3.29E-08 |

**Figure 2.**
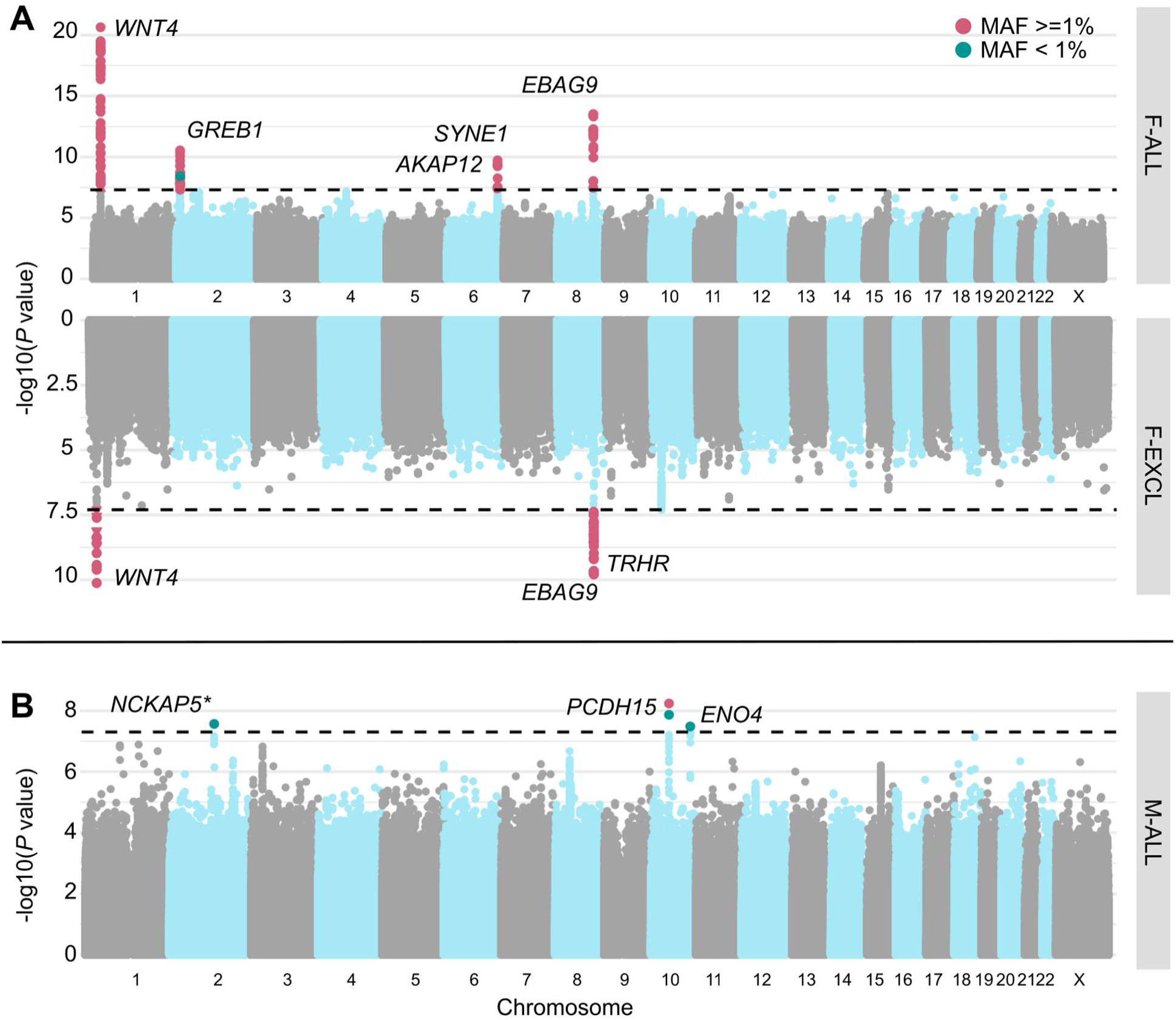
Miami and Manhattan plots for selected infertility meta-analyses. (A) Genetic variants associated with female infertility of all causes (F-ALL) (top) and idiopathic infertility (unknown causes) defined by exclusion of known causes such as anatomical or anovulatory causes, PCOS, endometriosis, or uterine leiomyomas (bottom). (B) Genetic variants associated with male infertility of all causes (M-ALL). Each point depicts a single SNP, with genome-wide significant (GWS) SNPs (P<5E-08, dashed line) coloured in pink for common variants with minor allele frequency (MAF)>=1% and green for those with MAF<1%. SNPs are annotated with the mapped gene. **\*** indicates that lead variant is reported in only one cohort.

Among the variants associated with multiple subtypes of female infertility is rs1964514, an intronic variant in *PKHD1L1* (OR (95% CI) for F-ALL=1.13 (1.09-1.16), F-EXCL=1.13 (1.09-1.17), F-INCL=1.18 (1.11-1.25)). This variant is 76 kb upstream of *EBAG9*, an oestrogen-responsive gene previously reported to have a recessive association with female infertility^28^ and thought to suppress maternal immune response during pregnancy^29,30^. We also identified an intronic variant in *WNT4*, rs61768001, associated with three categories of female infertility (F-ALL=0.909 (0.891-0.927), F-EXCL=0.923 (0.902-0.946), F-INCL=0.870 (0.839-0.903)). *WNT4* is highly pleiotropic for female reproductive traits, as it is reported to associate with gestational length^31^, uterine fibroids^32,33^, endometriosis^34,35^, female genital prolapse^27^, and bilateral oophorectomy^27^. Such pleiotropy is expected, as *WNT4* is a key regulator of female reproductive organ development in embryogenesis^36–38^.

The nearest gene to the idiopathic infertility-associated variant rs111597692 (F-EXCL OR=1.16 (1.10-1.22)) is *TRHR*, which encodes the thyrotropin-releasing hormone receptor. Mice with *TRHR* knockouts display a phenotype similar to primary ovarian insufficiency^39,40^. The F-ANOV associated variant rs72827480 (OR=0.905 (0.873-0.938)) colocalises with a testis-eQTL for *INHBB* in the GTEx Project^41^ (posterior probability (PP) of shared causal variant=91.6%) (Supp. Table 4). *INHBB* encodes the beta subunit of inhibin B, which regulates hypothalamic, pituitary, and gonadal hormone secretion^42^, and ovarian follicle and oocyte development^43^.

Finally, an intronic variant in *ENO4*, which is expressed in the testis and may play a role in sperm motility^44^, is associated with male infertility (rs139862664, OR=0.388 (0.277-0.543)). Male mice with *ENO4* knockouts display infertility, abnormal sperm morphology and physiology, and decreased testis weight, among other altered male reproductive tract phenotypes^45^.

#### Genetic relationships between infertility and female reproductive conditions

Genome-wide, we observed positive genetic correlation between endometriosis and F-ALL (*r*_g_ (SE)=0.585 (0.0785), *P*=8.98E-14) and F-INCL (*r*_g_=0.710 (0.115), *P=*5.94E-10). We also observed positive correlation between F-ANOV and PCOS, the most common cause of anovulatory infertility (*r*_g_=0.403 (0.131), *P*=2.20E-3), and negative correlation between F-ANOV and spontaneous dizygotic twinning, a heritable metric of female fecundity that captures the propensity for multiple ovulation^46^ (*r*_g_=-0.740 (0.182), *P*=4.93E-05).

Two loci associated with both endometriosis and female infertility - *WNT4* and *ESR1* - may share the same putative causal variant (PP>93.6%, Supp. Table 5). Variants in both these genes have previously been associated with endometriosis-related infertility^47–50^. While *GREB1* and *SYNE1* also contain overlapping signals for infertility and endometriosis, there is strong evidence against shared causal variants (PP>75%, Supp. Table 5). Finally, three of eight loci for anovulatory infertility - *INHBB*, *PITPNB*, and *CHEK2* - may share a causal variant with PCOS (PP>89.2%, Supp. Table 5).

### Selection pressure may explain the persistence of some infertility-associated variants in the population

The genome-wide SNP heritability estimates (on the liability scale, accounting for disease prevalence^54^) for all categories of infertility are <10% (lowest for M-ALL at 1.12% (SE=0.93) and highest for F-ANOV at 9.54% (2.16)) (Supp. Table 6). This is lower than heritability estimates of two-thirds of all heritable binary phenotypes in the UK Biobank with population prevalence similar to that of infertility (64 phenotypes with *Z*>4 and prevalence <5%)^53^. We hypothesised that infertility risk-increasing alleles are subject to negative selection^55^, so we tested whether there was evidence for: (i) variants associated with infertility in loci under historical or recent directional selection^56–58^, or (ii) recent directional selection (over the last 2,000 to 3,000 years) measured by singleton density scores (SDSs)^56^ and balancing selection measured by standardised BetaScan2 scores (StdB2)^59^ at infertility loci.

While we found no genome-wide signature of directional selection against infertility (Supp. Text), we observed extreme SDSs (in the highest 99.75^th^ percentile (%ile) of SNPs within 10kb of a GWAS Catalog variant) at the *EBAG9* locus associated with female infertility, indicating recent positive selection (Figure 4 and Supp. Table 7). *EBAG9* is associated with infectious response phenotypes, suggesting that the locus may be under selection for its effects on the immune system. We additionally observed signatures of balancing selection, which maintains multiple alleles in the population through mechanisms such as heterozygote advantage or time-varying fitness^60,61^, at the female infertility loci *GREB1* (StdB2 in the 98.6^th^-99.4^th^ %ile of SNPs within 10kb of a GWAS Catalog variant) and *INHBB* (98.5^th^ %ile), and the male infertility locus *PCDH15* (98.7^th^ %ile); however, variants at these loci with high probability of association with infertility did not have high balancing selection scores (Supp. Figure 2 and Supp. Table 7).

**Figure 3.**
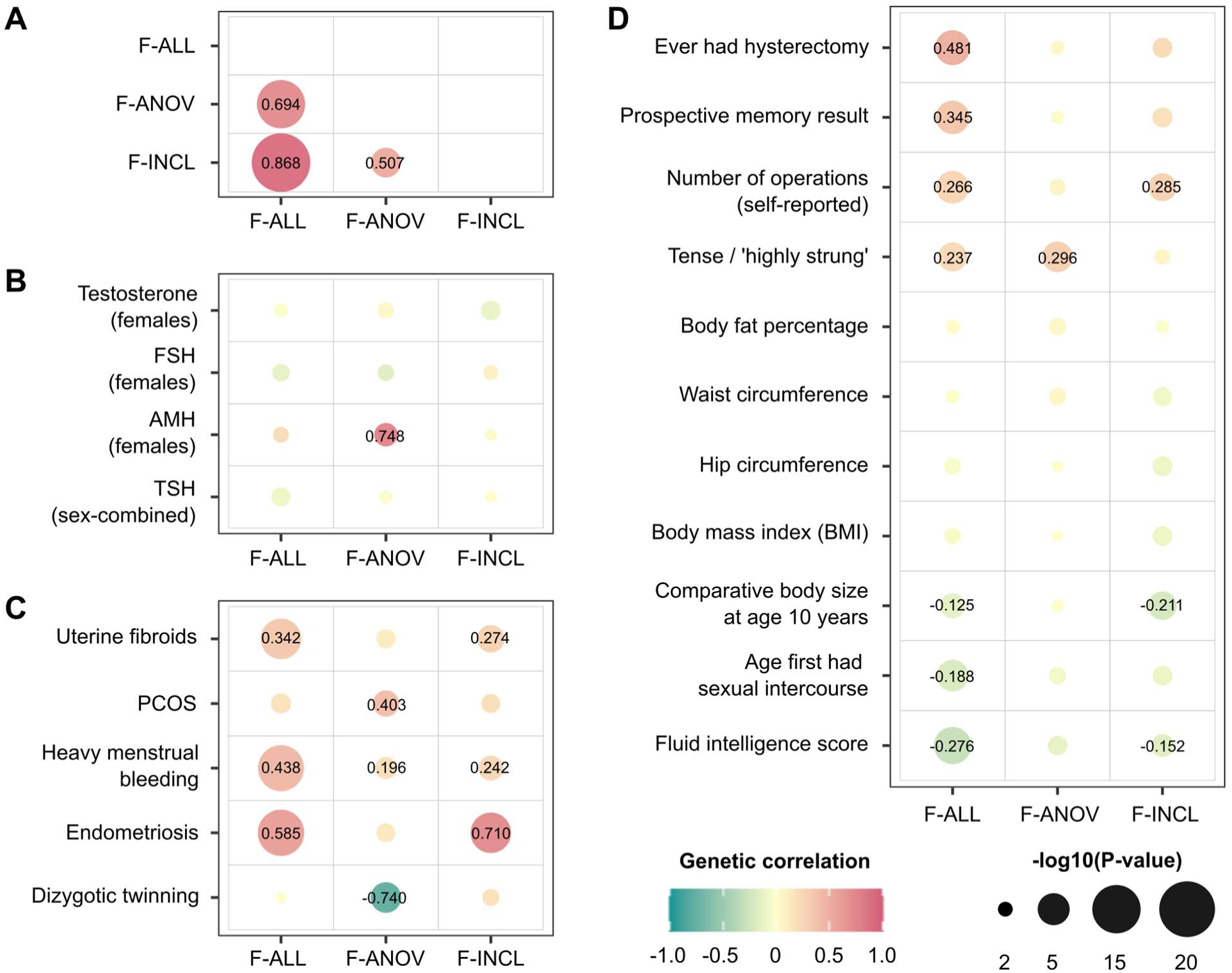
Genetic correlations between female infertility and other phenotypes. SNP-based genetic correlations (*r*_g_) between significantly heritable phenotypes (*Z*>4) were estimated using LD-score regression, performed using the LDSC software^51^ on a subset of 1 million HapMap3 SNPs^52^. Points are coloured by *r*_g_ estimate, scaled by significance (-log10(P)), and labelled with the associated *r*_g_ estimate if nominally significant without correction for multiple testing (*P*<0.05). (A) Genetic correlations among the three significantly heritable definitions of female infertility (all cause=F-ALL, anovulatory=F-ANOV, and idiopathic infertility defined by inclusion=F-INCL). (B) Genetic correlations between female infertility traits and reproductive hormones: testosterone, follicle stimulating hormone (FSH), and anti-Mullerian hormone (AMH, publicly available summary statistics) in female-specific analyses, and thyroid stimulating hormone (TSH, publicly available summary statistics) from sex-combined analysis. (C) Genetic correlations between female infertility traits and female reproductive conditions, with summary statistics generated from the largest available European-ancestry studies for each trait (see Methods). PCOS=polycystic ovary syndrome. (D) Genetic correlations between female infertility traits and selected heritable phenotypes (*Z*>4) in the UK Biobank, as generated by the Neale lab^53^. Correlations with all heritable phenotypes can be found in Supp. Table 12.

**Figure 4.**
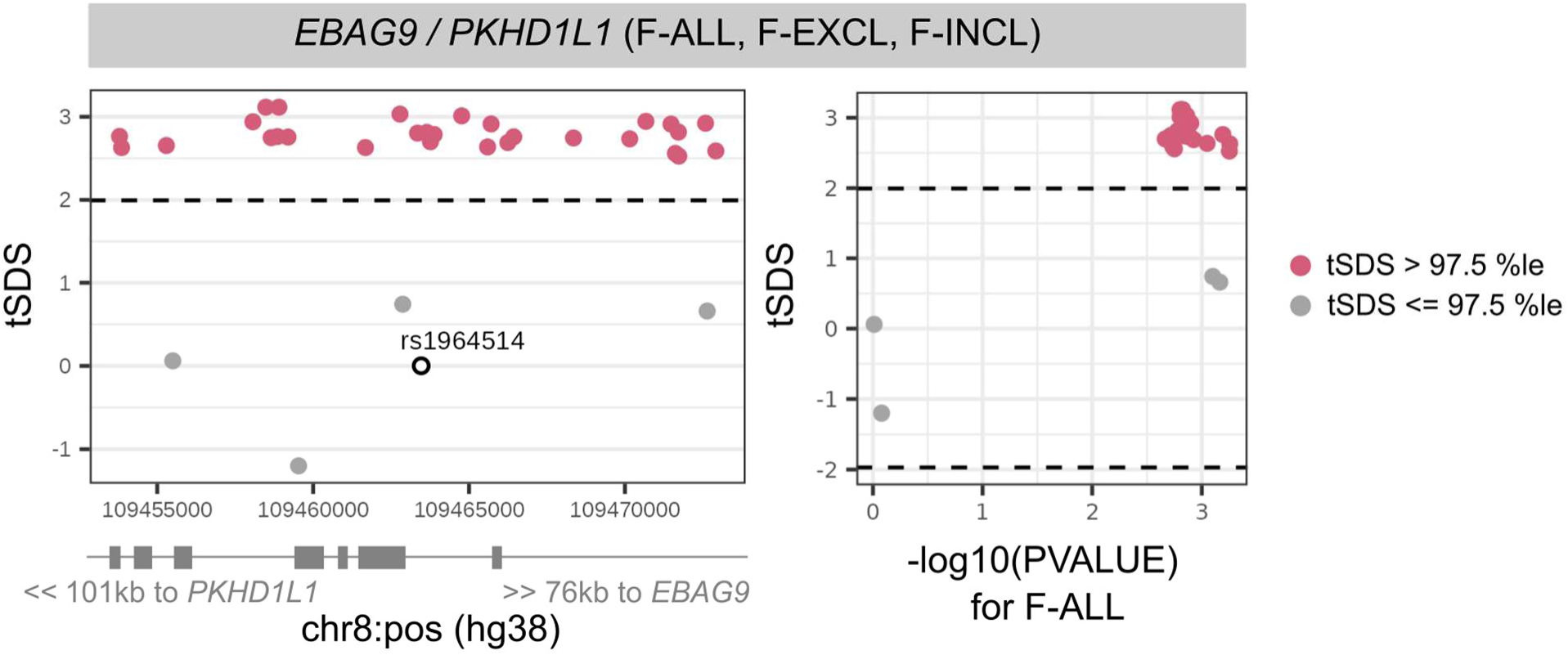
Directional selection scores at infertility-associated *EBAG9* locus. Recent directional selection, as measured by trait-aligned Singleton Density Scores (tSDSs) at the *EBAG9* locus. The window of +/- 10 kb around the lead variant associated with female infertility of all causes (F-ALL) is displayed, along with the location of nearest gene transcription start sites (TSSs). The tSDSs are aligned to the infertility-risk increasing allele, wherein a positive tSDS indicates positive selection for infertility-risk increasing allele at the locus. Dashed lines indicate 2.5th percentile (%ile) and 97.5th %ile of SDSs, and variants below or above this threshold respectively are coloured in pink. Left: Locus plots depicting genomic position on the x-axis and tSDS on the y-axis. The lead variant rs1964514 (open circle) is not present in the tSDS dataset and thus assigned a score of 0. Right: Scatter plots depicting relationship between -log10 of the GWAS p-value for the variant association with F-ALL on the x-axis and tSDS on the y-axis.

### Genetic determinants of reproductive hormone levels

#### Identification of novel reproductive hormone loci

As hormone dysregulation is central to many infertility diagnoses^18,19^, we conducted sex-specific GWAS meta-analyses of five reproductive hormones - follicle-stimulating hormone (FSH) (N_female_=57,890, N_male_=6,095), luteinising hormone (LH) (N_female_=47,986, N_male_=6,769), oestradiol (N_female_=97,887, N_male_=39,165), progesterone (N_female_=18,368), and total testosterone (N_female_=246,862, N_male_=243,951) - collected at assessment centre visits or identified through EHRs, in six cohorts and publicly available summary statistics (Supp. Table 9). We identified GWS loci associated with FSH (9 novel/2 previously known in females (F) and 0/1 in males (M)), LH (4/2 in F and 1/0 in M), oestradiol (1/1 in F and 2/4 in M), and testosterone (35/118 in F and 55/206 in M), but found no genetic variants associated with progesterone (Figure 5, Supp. Figure 3, and Supp. Figure 4). Several of the reported signals we replicated are near genes encoding the hormone-specific subunits themselves, such as *FSHB* for FSH and *LHB* for LH, or enzymes for steroid hormone metabolism, such as *CYP3A7* for oestradiol and *HSD17B13* for testosterone (Supp. Text).

**Figure 5.**
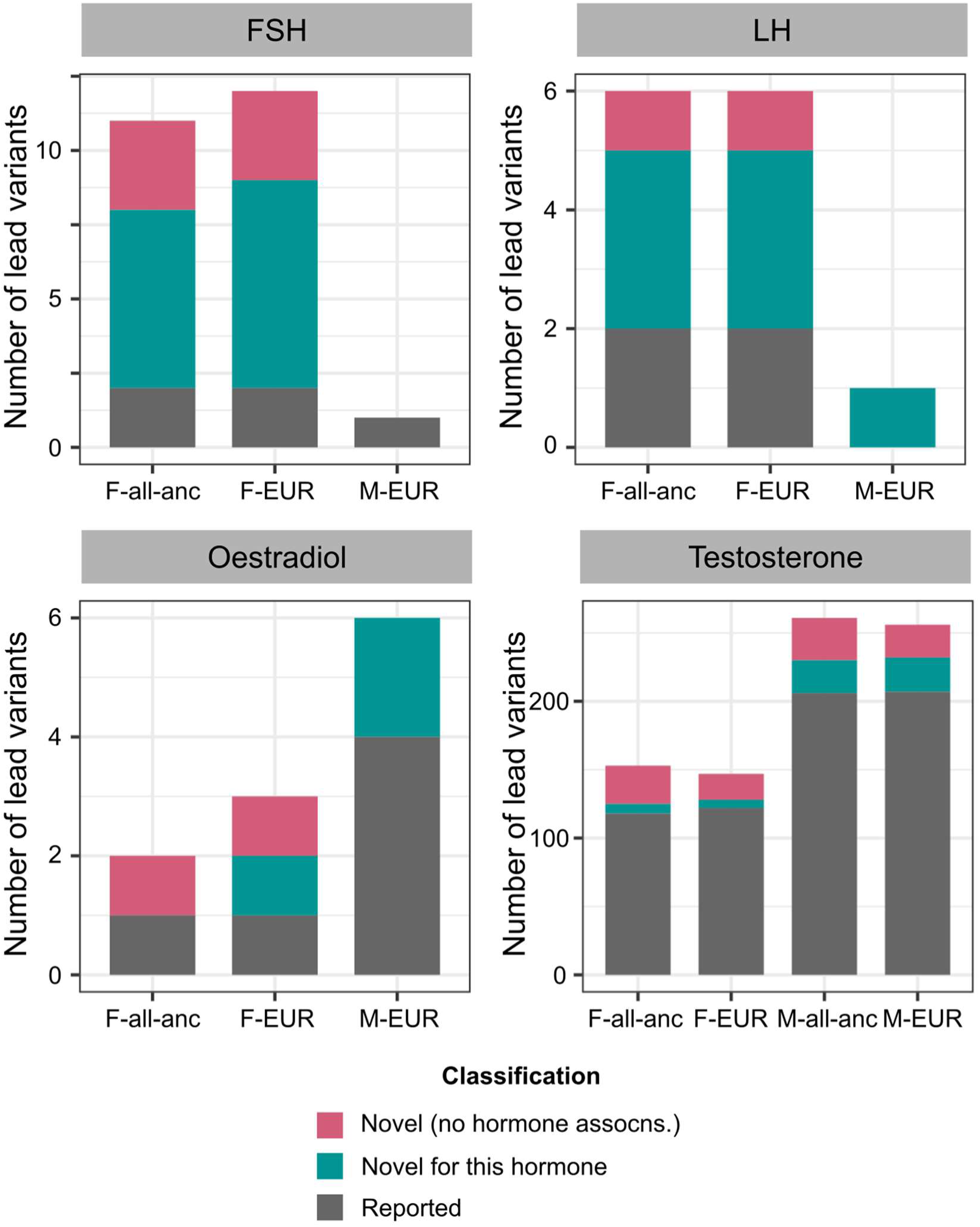
Number of novel and reported reproductive hormone associations. Each panel displays a different hormone (FSH=follicle-stimulating hormone, LH=luteinising hormone). Lead variants in each analysis stratum (F=female-specific, M=male-specific, all-anc=all ancestry meta-analysis, EUR=European-only meta-analysis) are classified as: (1) novel (no hormone associations) if they are not in LD (*r*^2^<0.1) with, and conditionally independent of (conditional *P*-value *P_cond_*<0.05), any variants within a 1Mb window of the lead variant that are associated with 28 reproductive hormones in the GWAS Catalog^62^, plotted in pink, (2) novel for this hormone if they are not in LD (*r*^2^<0.1) with, and conditionally independent of (*P_cond_*<0.05), the respective hormone-associated variants within a 1Mb window of the lead variant, plotted in green, and (3) reported otherwise, plotted in grey. Note the different Y-axis scales in each subplot. assocns.=associations.

We further classified lead variants as entirely novel hormone associations by defining a protocol based on linkage disequilibrium (LD) and conditional independence from published SNPs associated with any of 28 reproductive hormones in the GWAS Catalog^62^ (see Methods for detailed classification protocol).

We found 39 novel variants for testosterone in men, including those near *SPOCK1* (rs1073917: β (SE)=-0.0160 (0.0029), *P*=4.69E-08), which is a target for the androgen receptor^63^, *NR4A3* (rs10988865: β=-0.0161 (0.0029), *P*=4.33E-08), which coordinates the cellular response to corticotropin-hormone and thyrotropin-hormone releasing stimuli^64,65^ and regulates adipogenesis^66^, and obesity-associated genes *ANKS1B* (rs144998814: β=0.133 (0.0162), *P*=2.34E-16) and *ANO10* (rs6809522: β=0.016 (0.0029), *P*=3.00E-08)^67^ (Supp. Table 10). The 28 novel reproductive hormone variants associated with testosterone in women include those near *LAMTOR4* (rs17250196: β=-0.131 (0.0067), *P*=4.02E-86), associated with hyperthyroidism^39^ and age at menarche and menopause^68^, obesity-associated *CCDC146* (rs138240474: β=-0.116 (0.0207), *P*=2.03E-08)^67^, which is also expressed in the fallopian tubes and endometrium^69,70^, and *SLC8A1* (rs12611602: β=0.0163 (0.003), *P*=3.79E-08), which causes increased pancreatic beta cell proliferation and insulin secretion in knockout mouse models^71^. Finally, we report lead SNPs independent of previously published hormone variants in the *HELQ* locus for FSH-F (rs4235062: β=-0.046 (0.0065), *P*=1.50E-12), *TMEM150B* locus for FSH-F (rs28875253: β=-0.0599 (0.0061), *P*=9.90E-23) and LH-F (rs11668309: β=0.0519 (0.0071), *P*=3.91E-13), and in the *SOX15-SAT2* locus for oestradiol-F (rs3933469: β=0.0363 (0.0051), *P=*1.02E-12) (Supp. Table 10).

Our results were robust to the inclusion of summary statistics from publicly available datasets, and there was no evidence for heterogeneity in variant effects across cohorts (Supp. Text).

#### Sex-specific genetic architecture of testosterone

Only 9.80% (of 153 total) lead variants for testosterone in females and 5.75% (of 261 total) lead variants for testosterone in males reach GWS in both sexes; and 45.9% of variants have opposing directions of effect in men and women (Supp. Figure 6). Indeed, we found no significant genetic correlation between testosterone in men and women (*r*_g_ (SE)=0.0361 (0.0428), *P=*0.399). The heritability of testosterone in women is enriched in the adrenal gland (*P*=1.03E-03) and hepatocytes (*P*=9.36E-04); but only the latter is enriched for the heritability of testosterone in men (*P*=3.61E-04), as is the liver more broadly (*P*=1.16E-06) (Supp. Figure 10, stratified LD-score regression performed across 205 tissues and cell-types from the Genotype Tissue Expression (GTEx) Project database^41^ and the Franke lab single-cell database^72^). Finally, although testosterone regulates several traits hypothesised to be under sexual selection and may be under selection itself^73^, we do not find significant genome-wide directional selection for testosterone in men or women (mean genome-wide trait-SDS is not significantly different from 0, both *P*>0.05) (Supp. Text).

### Genetic relationships between female infertility, reproductive hormones, and obesity

We observed no genome-wide genetic correlations between any category of female infertility and: (i) any reproductive hormone in this study, or (ii) thyroid stimulating hormone (TSH), or (iii) anti-Mullerian hormone (AMH), the latter two based on publicly available summary statistics^74,75^ (all *P*>0.05, Figure 3B). Mendelian randomisation (MR) analyses indicated a genetically causal protective effect of FSH on risk of F-ALL (OR (95% CI)=0.776 (0.678-0.888), *P*=2.15E-04) and F-EXCL (0.716 (0.604-0.850), *P*=1.26E-04) (Supp. Table 11).

We found evidence for shared variants between hormones and infertility at the *FSHB* locus associated with FSH, LH, and testosterone (PP>84.8% for colocalisation with F-ANOV), and the *ARL14EP* locus associated with LH (PP=89.3% for colocalisation with F-ANOV) (Supp. Table 12). There was no evidence for colocalisation at any of the >300 other GWS loci associated with infertility or reproductive hormones in our study (Supp. Table 12). Our results suggest that while these traits are not significantly correlated at a genome-wide level, a small number of genes may drive infertility linked to hormone dysregulation.

Across 703 heritable phenotypes in the UK Biobank, we found 15 traits to be genetically correlated with female infertility, which we broadly group into: female reproductive conditions (such as having had a hysterectomy, *r*_g_ (SE)=0.481 (0.0963)), general illness (such as number of operations, *r*_g_=0.266 (0.0588)), and cognitive test results (overall prospective memory test *r*_g_=0.345 (0.0736), overall fluid intelligence *r*_g_=-0.276 (0.0502)) (Figure 3D and Supp. Table 13). 24 obesity-related traits, including body mass index (BMI), waist-to-hip ratio (WHR), and body fat percentage, are correlated with testosterone and FSH, but are not genetically correlated with any category of female infertility (all *P*>0.05, Figure 3D, Supp. Figure 7, and Supp. Table 13). However, MR analyses using genetic instruments for BMI, WHR, and WHR adjusted for BMI (WHRadjBMI)^67^ indicated evidence for bi-directional causal relationships between infertility and abdominal obesity independent of overall obesity. While genetically predicted WHRadjBMI is a risk factor for F-ALL (OR (95% CI)=1.10 (1.05-1.16), *P*=1.71E-04) and F-ANOV (1.29 (1.16-1.45), *P*=4.66E-06), the latter is itself causal for increased WHRadjBMI (β (SE)=0.0547 (0.0133), *P*=3.74E-05) (Supp. Table 11).

Variants associated with all-cause female infertility are in genes enriched for expression in ovarian stromal cells (partitioned heritability *P*=2.52E-03). We did not find significant enrichment of infertility heritability in any of the 205 tissues and cell-types from the GTEx project database^41^ and the Franke lab single-cell database^72^.

### Rare variant contribution to reproductive-hormone and infertility genetics

We analysed the 450k UK Biobank exome sequencing dataset to characterise the association between rare coding variation (MAF<1%) and binary traits with >100 cases (F-ALL (3,746 cases, 260,413 controls), F-EXCL (3,012 cases, 261,147 controls), and M-ALL (650 cases, 222,393 controls)), and quantitative traits with >10,000 participants (FSH-F (N=20,800), LH-F (N=16,391), oestradiol-F (N=54,609), and testosterone (N_female_=197,038, N_male_=197,340) (Figure 1)). Gene-burden analyses implicate the *PLEKHG4* gene, which is highly expressed in the testis and ovary, for F-EXCL (burden test OR (95% CI)=1.04 (1.02-1.06) when aggregated across all variant annotations with MAF<1%, Cauchy *P*=1.37E-08) (Supp. Table 14). Rare variants in *PLEKHG4* cause cerebellar ataxia^76^, which is a feature of some syndromes that also cause steroid hormone deficiency and hypogonadism^77,78^.

#### Novel genes for testosterone implicated by gene burden analyses

Gene-based analyses identify 27 genes associated with testosterone-F and 24 genes for testosterone-M (*P*<5E-06), of which eleven have not previously been implicated in GWASs (Supp. Text). We report the first known association of *HSD11B1* with testosterone-F (burden test *P*=1.93E-06 when aggregated across missense variants with MAF<0.01%); pathogenic variants in this gene are reported to cause hyperandrogenism due to cortisone reductase deficiency^79,80^ (Supp. Figure 11 and Supp. Table 14). We also report the association of testosterone-M with *HSD17B2* (burden test *P*=1.33E-11 when aggregated across pLoF variants with MAF<0.1%), which encodes the enzyme hydroxysteroid 17β-dehydrogenase 2 that catalyses the oxidation of oestradiol, testosterone, and dihydrotestosterone to less active forms and thus regulates the biological potency of steroid hormones^81,82^ (Supp. Figure 11 and Supp. Table 14).

#### Increased risk of infertility in individuals carrying rare testosterone-associated variants

Two genes associated with testosterone in female UK Biobank participants are also associated with infertility risk (*P*<1.00E-03, Bonferroni adjustment for 50 unique genes): *TRIM4* (F-ALL, burden test OR=1.03 (1.01-1.05), *P*=4.05E-04 across all variants with MAF<0.1%) and *CYP3A43* (F-EXCL, burden test OR=1.02 (1.01-1.03), *P*=4.84E-04 across all variants with MAF<1%). The latter encodes the steroid hormone metabolic enzyme testosterone 6-beta-hydroxylase; but neither gene has previously been implicated in infertility.

Finally, we identified 29 unique genes carrying rare variants (MAF<1%) associated with testosterone in male or female participants in the UK Biobank. Eighteen of the 29 genes also contain common testosterone-associated variants from GWASs (MAF>1%), but the rare variant has a larger absolute effect size in the majority (83%) of these (Figure 6A, Supp. Table 15, and Supp. Text).

**Figure 6.**
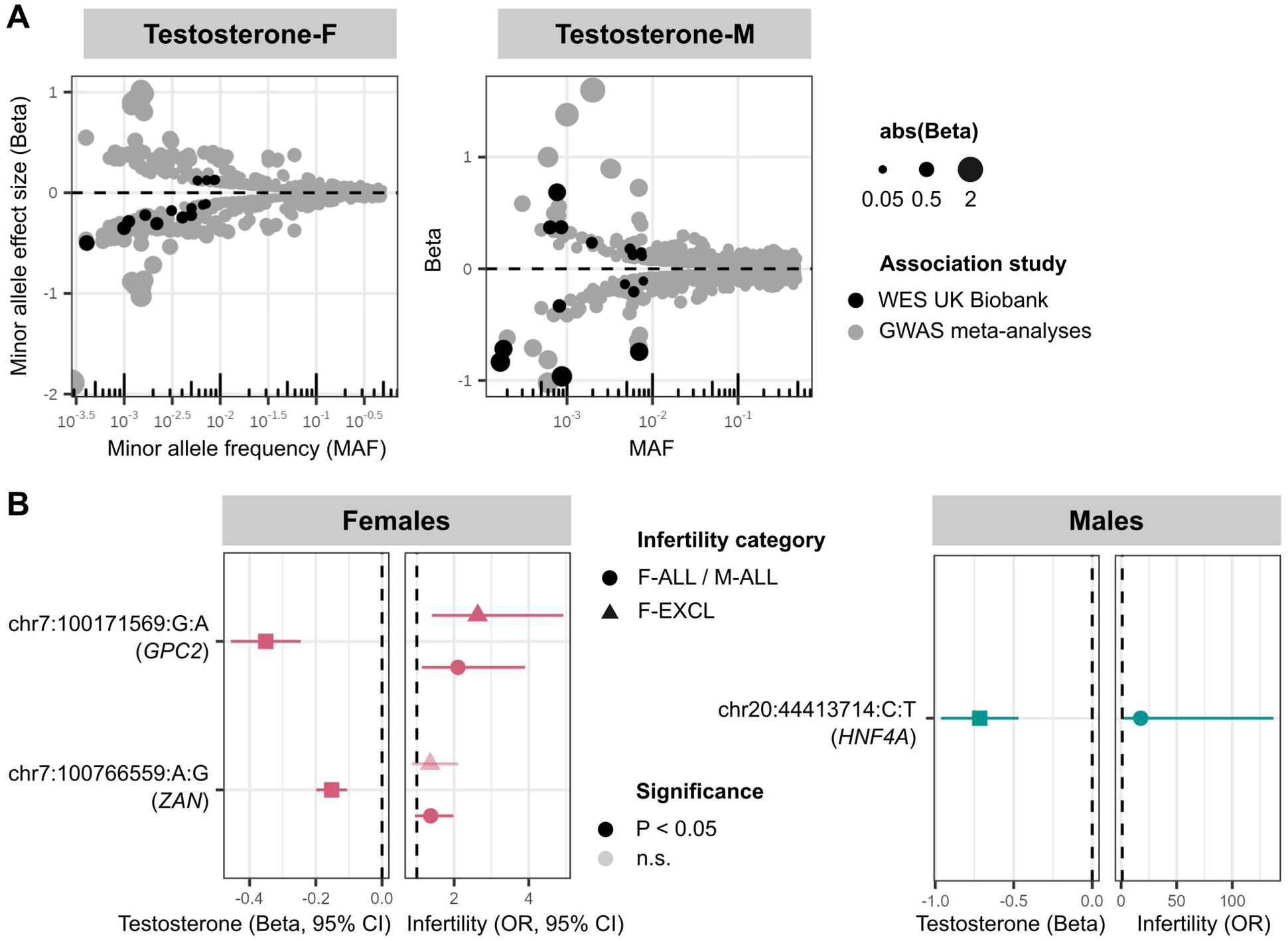
Rare variants associated with testosterone and infertility in UK Biobank whole exome sequencing (WES) analyses. (A) Effect size versus allele frequency of genetic variants associated with total testosterone. Variants discovered at genome-wide significance (*P*<5E-08) in GWAS meta-analyses (coloured in grey) and exome-wide significance in the UK Biobank WES analyses (coloured in black) are plotted, sized by the absolute value of their effect size. Effect sizes are aligned to the minor allele, plotted against MAF on the log x-axis. (B) Effects of testosterone-associated rare variants on infertility in females (left) and males (right). Per gene, the variant with lowest *P*-value of all variants that reach exome-wide significance (*P*<1E-07) in UK Biobank WES analyses for testosterone is displayed, for all variants with nominally significant effects on infertility. Effect sizes (β and 95% confidence intervals (CIs) for the variant effect on testosterone are to the left of each plot, and effect sizes (odds ratios (ORs) and 95% CIs) for the variant effect on infertility are to the right of each plot. Variants that reach nominal significance (*P*<0.05) are coloured in solid shapes.

The eleven novel testosterone associations include a female testosterone-lowering missense variant in *STAG3* (chr7:100204708:C:T, β=-0.284, *P=*2.31E-08); *STAG3* is also associated with primary ovarian insufficiency in women^83,84^, and induces female infertility through the absence of oocytes in knockout mouse models^39^. While we did not find significant association between the *STAG3* variant and female infertility in the UK Biobank (*P*>0.05), we observed increased risk of idiopathic infertility in women carrying a novel testosterone-lowering variant in *GPC2* (chr7:100171569:G:A, F-EXCL OR=2.63 (1.40-4.92), *P*=1.25E-03) (Figure 6B). *GPC2* is highly expressed in the testis, and *GPC2-*knockout mouse models display reduced adrenal gland size^39^.

The gene has not previously been reported to be associated with testosterone or infertility. Taken together, our results indicate a potential role for infertility driven by rare hormone-disrupting variants.

## Discussion

Our large-scale genetic investigation of infertility and related reproductive phenotypes in over 1 million individuals identified 19 genetic loci associated with female infertility, two with male infertility, and novel variants for the reproductive hormones FSH (3 novel variants), LH (1), oestradiol (1), and total testosterone (28) in women and for total testosterone in men (39). Through rare-variant and gene-based analyses in the UK Biobank, we additionally identified *PLEKHG4* associated with female infertility and 50 genes for testosterone, including the first reported hormone-associated variants in some members of the hydroxysteroid dehydrogenase enzyme family. We found evidence at non-hormonal, pleiotropic, infertility loci for recent directional selection (*EBAG9*) and balancing selection (*GREB1, INHBB*, *PCDH15*). Although there was evidence for distinct genetic architectures of infertility and reproductive hormones, we showed that individual genes containing rare protein-coding variants associated with testosterone (*GPC2*, *CYP3A43*, *TRIM4*) were also associated with higher risk of infertility in the UK Biobank.

Previous efforts to catalogue the genome-wide architecture of infertility have relied on proxy measures such as childlessness and number of children ever born^24,25^, which may be confounded by behavioural, socio-economic, and lifestyle factors. While we did find modest genetic correlation between female infertility and age at first sexual intercourse (-18.8%), indicating that the latter captures some shared biology with fertility, our meta-analyses did not replicate the associations of infertility proxy variables with putative behavioural loci for risk-taking^85,86^ or educational attainment^85,87–89^. Instead, we nominate genes with putative roles in both male and female gonads, such as *TRHR* for ovarian insufficiency^39,40^ and *ENO4* for sperm motility^44^.

The strong genetic correlation of 71% between idiopathic infertility and endometriosis may indicate that some proportion of idiopathic cases are due to under-diagnosis of endometriosis, whose early treatment may prevent future infertility^15,90^. Our subtype-specific analyses highlight the value in dissecting heterogeneous causes of infertility. For example, PCOS is a heritable cause of up to 80% of anovulatory infertility cases that may be treated through induced ovulation^17,91,92^. However, as only three of eight loci for anovulatory infertility colocalise with known PCOS signals and the genetic correlation between these traits is only 40%, other hypothalamic-pituitary-ovarian disorders, endocrinopathies (hypothyroidism, hyperprolactinaemia, etc.) and ovarian insufficiency may also contribute significantly to the genetic aetiology of anovulatory infertility and require treatments different from those for PCOS-associated infertility^93^. Weight loss for overweight patients is often recommended as beneficial for fertility^94,95^, but we did not find substantial genetic correlation between obesity and infertility. Our findings add genetic support to evidence from randomised controlled trials demonstrating no fertility benefits from short-term weight loss in overweight and obese women^96^. Instead, we observed bi-directional causal relationships between abdominal obesity and anovulatory infertility, suggesting physiological feedback mechanisms whose complex interplay requires deeper study. Taken together, these results suggest a critical need for a richer understanding of the genetic and non-genetic contributions to infertility.

The testes and ovaries were not significantly enriched for the heritability of infertility or testosterone, despite being reproductive organs that are major sites for testosterone production^97,98^. However, neither organ is disaggregated into tissues or cell types in the GTEx database, so gene expression profiles may not capture cell-type specific effects. Indeed, we found enrichment of testosterone heritability in the androgen-secreting thecal cells and androgen-responsive granulosa cells of the ovary^99–101^, and female infertility in ovarian stromal cells. Although there are several causal roles hypothesised for stromal dysfunction in infertility, such as impaired folliculogenesis^102^, restricted blood flow^103^, and ovarian scarring^104^, more work is needed to robustly replicate these findings. In general, more functional studies of gonadal cell types, in both men and women, are needed to enable a mechanistic understanding of the genetic variation associated with reproductive hormones and infertility.

We employed a broad search strategy to maximise sample sizes for cases of infertility and reproductive hormone levels in our meta-analyses. Diagnostic criteria for infertility vary by country and have changed over time^1^, which may explain the wide spread in the prevalence of infertility across cohorts. Reproductive hormone values in this study were assayed using different methodologies, in primary care or hospital EHRs, and at different ages and stages of the menstrual cycle in women. A majority of samples in our study were derived from the UK Biobank and measured during and post-menopause (ages 40-69), whereas infertility occurs pre-menopause, so we urge caution in interpreting the lack of correlation between these traits. Although we were able to adjust for covariates such as age, which can account for some of the effect of menopause on hormone levels, we did not have the data granularity to account for hormonal fluctuations during the menstrual cycle and pregnancy. In the future, longitudinal GWASs that can incorporate mean and variance of hormone levels over the menstrual cycle, or phenotypes that calculate ratios between various hormones over time, will likely reveal fundamental biology that is missed by the broad-stroke assessments in this study.

Our results indicate that balancing selection and recent positive selection at pleiotropic loci may explain the persistence of genetic factors for infertility. For example, the *EBAG9* locus associated with female infertility is under directional selection, perhaps because *EBAG9*, which is highly expressed in CD34-/CD41+/CD42+ megakaryocytes^69,70^, plays a role in T-cell mediated cytotoxicity as part of the adaptive immune memory response to infection^105^. However, a complementary role for *EBAG9* may be in the placenta during early pregnancy, where reduction of *EBAG9* levels is associated with inappropriate activation of the maternal immune system and results in foetal rejection^106^.

In conclusion, in this comprehensive large-scale investigation of the genetic determinants of infertility and reproductive hormones across men and women, we identified several genes associated with infertility and analysed their effects on reproductive disease and selection pressures. We did not find evidence that reproductive hormone dysregulation and obesity are strongly correlated with infertility at the population level, but instead nominate individual hormone-associated genes with effects on fertility. Other genetic and non-genetic avenues must be explored to treat complex and heterogeneous fertility disorders that impact the physical, emotional, and financial well-being of millions of individuals across the globe.

## Methods

### Study populations and phenotype identification

#### Binary traits (infertility)

Cases were identified in UK Biobank, Copenhagen Hospital Biobank and Danish Blood Donor Study, deCode, Estonian Biobank, FinnGen, and Genes and Health (Supp. Text). We defined five categories of female infertility: all causes (F-ALL), anovulatory (F-ANOV), anatomical (F-ANAT, including tubal, uterine, and cervical origins), idiopathic infertility by exclusion of known causes (anatomical and anovulatory infertility, PCOS, endometriosis, and uterine leiomyoma) (F-EXCL), and idiopathic infertility by inclusion of a diagnosis code for idiopathic infertility (F-INCL), and male infertility of all causes (M-ALL). Cases were identified through self-report (F-ALL, F-EXCL, M-ALL) and through primary- and secondary-care codes (Supp. Table 1). Within each subtype, sex-matched controls were defined as individuals not identified as cases for that subtype.

#### Quantitative traits (reproductive hormones)

Hormones were included from UK Biobank, Avon Longitudinal Study of Parents and Children (ALSPAC), deCode, Estonian Biobank, and Genes and Health (Supp. Text). We extracted measurements of FSH, LH, oestradiol, progesterone, and testosterone from biobank assessment centres or primary- and secondary-care records (Supp. Table 16). If repeated measurements were available for an individual, we retained the recorded hormone value closest to the individual’s median hormone value over time. Each hormone was regressed on *age*, *age*^2^, and cohort-specific covariates specified below; the residuals from this regression were rank-based inverse normally transformed (RINTed) prior to GWAS.

### Meta-analysis of GWAS summary statistics

#### Genome-wide association testing

Association analyses were performed separately within each ancestry and sex stratum for all strata with at least 100 cases (binary traits) or 1,000 individuals (quantitative traits). For binary traits, each variant passing QC was tested for association under an additive model using REGENIE^107^ or SAIGE^108^, with adjustments for *age, age*^2^, and cohort-specific covariates, with the Firth correction applied to control for inflation at rare variants and traits with low case-control ratios^107,108^. For quantitative traits, the RINTed hormone value was tested for association under an additive model using REGENIE^107^ or SAIGE^108^, with adjustments for cohort-specific genetic covariates. Any deviations from this GWAS protocol are noted in the Supplementary Text.

#### Meta-analysis

Prior to meta-analysis, summary statistics from all studies underwent thorough quality control to retain variants that met the following criteria: (1) on the autosomes or X chromosome, (2) with imputation information score >0.8 (where available), (3) bi-allelic variants with A, C, G, T alleles, (4) with standard errors <10 and *P*-values in [0,1], and (5) without duplicate entries. Fixed-effects inverse-variance weighted meta-analysis was performed using METAL^109^. We report results from European-ancestry and all-ancestry meta-analyses for each trait. Genome-wide significance was established at *P*<5E-08.

### Identification and classification of lead variants

Distance-based pruning was used to identify lead variants as the SNP with the lowest *P*-value within each 1Mb window at all loci with at least one GWS variant with *P<*5E-08.

Hormone-associated variants were classified based on conditional analysis as (1) previously reported for the hormone of interest, (2) previously reported for any of 28 reproductive hormones, or (3) novel, based on SNP associations published in the GWAS Catalog as of 27 March 2023^62^ (Supp. Table 17). We adapted criteria developed by Benonisdottir *et al.* (2016)^110^ to classify novel variants as those that are not in LD with (*r*^2^<0.1), and conditionally independent of (*P_conditional_*<0.05), all published hormone-associated variants within 1 Mb; all other variants are considered to be previously reported. Conditional analysis was performed in GCTA-COJO^111^, with LD information for European-ancestry individuals derived from the 1000 Genomes dataset^112^.

For lead variants on the X chromosome and those from multi-ancestry analyses, for which estimating LD is more difficult due to differences in recombination rates and selection pressures between sexes and populations^113–115^, we did not use the above LD-based classification system. Instead, a lead SNP was considered novel if it was not within 1 Mb of a published hormone-associated variant or if its effect was independent of published variants within a 1 Mb window (*P_conditional_*<0.05), and reported if not.

### SNP-based heritability

The following analyses, which rely on population-specific LD patterns, were restricted to European-ancestry summary statistics with pre-computed LD-scores based on European-ancestry individuals in the 1000 Genomes dataset^112^, restricted to HapMap3 SNPs^52^. We estimated the SNP-based heritability (*h_G2_*) of a trait from GWAS summary statistics using LD-score regression as implemented in the LDSC software^51^. For infertility traits, the observed-scale heritability (*h_obs2_*) was converted to liability-scale heritability (*h_liab2_*), which accounts for the disease prevalence in the sample (*k*) and population (*K*), under the assumption that sample prevalence equals the population prevalence^54^.

### Genetic correlations

LDSC was used to estimate genetic correlations between infertility traits, hormone levels, and a collection of other phenotypes in the UK Biobank in European-ancestry individuals. To simplify computation of *r*_g_ across a large number of traits, we used an extension of the LDSC software which allows for simultaneous estimation of multiple genetic correlations^116^.

We estimated genetic correlations among the three categories of female infertility with significant heritability (*Z*>4)^51^: F-ALL, F-ANOV, and F-INCL, as well as among heritable female reproductive hormones (FSH and testosterone in females). We additionally obtained summary statistics from GWASs of thyroid stimulating hormone (TSH)^75^ (sex-combined analysis, N=247,107 participants) and anti-Mullerian hormone (N=7,049 pre-menopausal participants)^74^ from the largest publicly available European-ancestry studies to date. We also tested for genetic correlations between infertility and reproductive hormones. Significant *r*_g_ after multiple testing was established at 2.38E-03 (FWER controlled at 5% across 21 tests using the Bonferroni method).

We collated European-ancestry GWAS summary statistics for four female reproductive disorders: (1) endometriosis from Rahmioglu *et al.* (2023)^35^, (57,248 cases and 698,764 controls), (2) heavy menstrual bleeding by meta-analysing GWAS data from Gallagher *et al.* (2019)^117^ and FinnGen data freeze 9^27^ (31,309 cases and 318,510 controls), (3) PCOS by meta-analysing GWAS data from Tyrmi *et al.* (2022)^92^ and a UKBB-based GWAS (14,467 cases and 430,267 controls), and (4) uterine fibroids by meta-analysing GWAS data generated by the Neale lab^53^ and FinnGen data freeze 9 ^27^, (42,446 cases and 588,955 controls). We additionally obtained summary statistics from a GWAS of spontaneous dizygotic (DZ) twinning (8,265 cases (mothers of DZ twins) and 264,567 controls; plus 26252 DZ twins and 417,433 additional controls) from Mbarek *et al.* (2024), the largest European-ancestry study of female fecundity to date^46^. Significant *r*_g_ after multiple testing was established at 2.00E-03 (FWER controlled at 5% across 25 tests using the Bonferroni method).

We downloaded LD-score formatted summary statistics for European-ancestry individuals across 703 heritable phenotypes (*Z*>4) from the Neale lab round 2 collection^53^. The number of effectively independent phenotypes estimated by the Neale lab (*M_eff_*=340) was used to establish significant *r*_g_ after multiple testing at 2.45E-05 (FWER controlled at 5% across 2,040 tests using the Bonferroni method).

### Mendelian randomisation

The following analyses were all performed with summary statistics from European-ancestry GWASs, using the TwoSampleMR v0.5.7 package^118^.

We constructed genetic instruments for BMI, WHR, and WHRadjBMI with female-specific lead variants from a recent European-ancestry GWAS meta-analysis with a maximum sample size of 434,785 female participants^67^. SNPs were weighted by their female-specific effect sizes. The mean F-statistic across all SNPs in each instrument indicated sufficient strength for MR (BMI=61.3, WHR=74.8, WHRadjBMI=84.7, recommended>10^119^). As the instrument GWASs included participants from UK Biobank, we conducted a sensitivity analysis to avoid bias from sample overlap between instrument and outcome GWASs by constructing obesity-trait instruments from an earlier release of summary statistics from the GIANT Consortium without UKBB participants^120^ (Supp. Table 11). As the WHRadjBMI instrument may be confounded due to adjustment for a correlated variable^121^, i.e. adjustment for BMI in the WHR GWAS, we performed multivariable MR with a joint instrument for BMI and WHR to estimate the BMI-adjusted causal effect of WHR on reproductive outcomes. We found no difference in effect estimates from MR conducted using an instrument for WHRadjBMI and multivariable MR (Supp. Table 19).

Hormone instruments were constructed for reproductive hormones in this study with F-statistic>10 (FSH-F=38.7, testosterone-F=66.1), using GWAS summary statistics from European-ancestry GWASs excluding UK Biobank participants to avoid sample overlap with outcome GWASs.

We also performed reciprocal MR to test the genetically predicted causal effects of infertility on obesity and reproductive hormone levels. Genetic instruments were constructed for subtypes of infertility with F-statistic>10 (F-ALL=51.0, F-ANOV=36.2), using GWAS summary statistics from European-ancestry GWASs excluding UK Biobank participants to avoid sample overlap with outcome GWASs. We assessed the causal direction between each pair of traits tested with Steiger filtering of instruments and the Steiger directionality test.

We report results from the inverse-variance weighted (IVW) method, the MR-Egger method which is robust to horizontal pleiotropy^122^, and the weighted median method which protects against outlier variants^123^ (Supp Table 11).

### Colocalisation

The following analyses were all performed with summary statistics from European-ancestry GWASs, using the Bayesian framework implemented in the coloc v5.1.0 package^124^ under a single causal variant assumption^125^. Only common variants (MAF>1%) within windows of +/- 50 kb around each lead variant for an infertility or reproductive hormone trait were retained. For each pair of traits tested for colocalisation, we set the prior probabilities of variants in a locus being causally associated with trait 1 (p_1_) and trait 2 (p_2_) to 1E-04 (99% confidence in a true association), and the prior for joint association p_12_ to 1E-06 (assuming equal likelihood of shared and non-shared causal variants for each trait in a locus) as recommended by the developers of coloc^125^. We tested five hypotheses: H0=no association with either trait in region, H1=association with trait 1 in region, but not trait 2, H2=association with trait 2 in region, but not trait 1, H3=association with both traits in region, but different causal variants, and H4=association with both traits in region, and a shared causal variant. A pair of traits were considered to colocalise if posterior probability of H4>50% and the ratio of posterior probabilities of H4/H3>5^124,126^.

We tested for colocalisation between each female infertility category and each female-specific hormone (FSH, LH, oestradiol, and testosterone) at all genetic loci associated with at least one of the pair of traits tested. The single male infertility locus with common variants (MAF>1%) in the European-ancestry analysis did not contain enough significant associations (only 12 common variants with *P*<1E-06) for colocalisation analyses.

Because we noticed that some lead variants for female infertility had previously been reported as associated with endometriosis and PCOS, we estimated the posterior probability (PP) of colocalisation of genetic signals between each category of female infertility and each of these two reproductive disorders. European-ancestry summary statistics for endometriosis and PCOS were obtained as described in the genetic correlations section above.

We assessed colocalisation of genetic signals for female infertility with eQTLs for all proximal genes with transcription start sites (TSSs) within 1 Mb of an infertility lead variant. Publicly available eQTL data was downloaded from the GTEx project^41^.

### Tissue and cell-type prioritisation

We estimated the polygenic contributions of genes with tissue-specific expression profiles to the heritability of infertility and hormones using stratified LD-score regression (partitioned heritability analyses)^51^. We restricted these analyses to traits with highly significant heritability in European-ancestry analyses (*Z*>7) (F-ALL, testosterone-F, and testosterone-M), as recommended by the developers, Finucane *et al.* (2015)^127^.

Gene sets and LD scores for 205 tissues and cell-types from the GTEx Project database^41^ and the Franke lab single-cell database^72^ were downloaded from Finucane *et al.* (2018)^128^. We established tissue-wide significance at -log10(*P*)>2.75, which corresponds to FDR<5%.

#### Ovarian cell types

As the ovary, a reproductive tissue of interest, is not well characterised in the GTEx project, we identified two publicly available single-cell gene expression datasets for ovarian cell types: (1) from Fan *et al.* (2019), who performed single-cell RNA sequencing on ovarian tissue from five adult women undergoing fertility preservation procedures with 20,676 cells across 19 identified cell types^129^, and (2) from Jin *et al.* (2022), who performed single-nucleus RNA sequencing on autopsy samples from four women (aged 49-54 years, with normal ovarian histology) with 42,568 cells across 8 identified cell types^130^. The datasets were aligned and filtered using the QC pipelines provided by the authors of each study, and clustered with identical parameters to replicate the results of each individual study. Gene sets for each cluster were identified as recommended by Finucane *et al.* (2018)^128^ - briefly, we identified differential expression between the cells in each cluster and all other clusters by using the Wilcoxon rank sum test implemented in Seurat v3.0^131–133^, and returned the top 10% of genes that are specifically expressed in each cluster (positive average log-fold-change values), ranked by differential expression *P*-value. We computed annotation-specific LD scores for these gene sets using hg38 coordinates for gene TSSs and TESs obtained from Ensembl^134^, across 1 million HapMap3 variants^52^ with LD information from European-ancestry individuals in the 1000 Genomes phase 3 dataset^112^.

### Overlaps with genetic regions under selection

To avoid confounding by population stratification, selection look-ups were restricted to GWAS summary statistics from European-ancestry individuals.

#### Directional selection

Following guidelines described by Mathieson *et al.* (2023)^25^, we identified 54 genomic regions under directional selection from three previously reported genome-wide scans: (1) 39 regions from the Composite of Multiple Signals (CMS) test, which infers historical selection on the order of the past 50,000 years^58^, (2) 12 regions from an ancient DNA scan that uses inferences of allele frequency from ancient genomes to determine selection over the past 10,000 years^57^, and (3) three regions from Singleton Density Scores (SDSs), which use the pattern of singleton variants to identify recent selection in the past 2,000 to 3,000 years^56^. For each genomic window under directional selection, we report the infertility-associated variants with the lowest *P*-value.

#### Singleton density scores

We downloaded publicly available SDSs for SNPs in the UK10K dataset^56^ to report the highest SDS (positive selection of derived allele over ancestral allele in the past 2,000 to 3,000 years) and lowest SDS (negative selection) within the +/-10kb window around each infertility or hormone lead SNP. To calculate trait-SDS for each phenotype, we aligned each SDS to the trait-increasing allele rather than the derived allele^56^. For each lead variant window containing variants with extreme SDSs (top 97.5th %ile or bottom 2.5th %ile), we report the direction of selection with respect to the trait-increasing allele. Percentiles of SDSs were evaluated only on a subset of variants within 10kb of any variant reported in the GWAS Catalog to account for genomic context. Further, as variants that are sub-GWS for a trait may nonetheless be under selection, we calculated the genome-wide mean trait-SDS in each bin of 1000 variants, ranked by *P*-value for the trait association, following the protocol outlined by Field *et al.* (2016)^56^.

#### Balancing selection

We accessed publicly available standardised BetaScan2 scores, which detect balancing selection using polymorphism and substitution data, for all SNPs in the 1000 Genomes dataset^59^. We tested whether the +/-10kb window around each infertility or hormone lead variant contained SNPs with scores in the 99th %ile of standardised BetaScan2 scores. Percentiles of SDSs were evaluated only on a subset of variants within 10kb of any variant reported in the GWAS Catalog to account for genomic context. For each lead variant window, we report the highest standardised BetaScan2 score and its percentile.

### Whole exome sequencing analyses in the UK Biobank

#### Exome sequencing quality control

##### Quality control outline

We first considered an initial set of “high quality” variants to evaluate the mean call rate and depth of coverage for each sample. We then ran a sample and variant level pre-filtering step and calculated sample-level QC metrics. Using these metrics, we removed sample outliers based on median absolute deviation (MAD) thresholds, and excluded sites which did not pass variant QC according to Karzcewski *et al*. (2022)^135^. We then applied a genotype-level filter using genotype quality (GQ), depth (DP), and heterozygote allele balance (AB). The resultant high-quality European call set consisted of 402,375 samples and 25,229,669 variants. For details see Supplementary Text.

#### Variant annotation

We annotated variants using Variant Effect Predictor (VEP) v105 (corresponding to gencode v39)^136^ with the LOFTEE v1.04_GRCh38^137^ and dbNSFP^138^ plugins, annotating variants with CADD v1.6^139^, and REVEL using dbNSFP4.3^140^ and loss of function confidence using LOFTEE. Complete instructions and code for this step are provided in our VEP_105_LOFTEE repository^141^, which contains a docker/singularity container to ensure reproducibility of annotations. We then ran SpliceAI v1.3^142^ using the gencode v39 gene annotation file to ensure alignment between VEP and SpliceAI transcript annotations. We defined ‘canonical’ transcripts to be used for variant-specific annotations as follows: set MANE Select^143^ as the canonical, where available, and if a MANE Select transcript is not present, set canonical and restrict to protein coding genes. Note that for VEP 105, this is equivalent to selecting the ‘canonical’ transcript in protein coding genes. Then, using the collection of missense, pLoF, splice metrics, and annotations of variant consequence on the ‘canonical’ transcript, we determine a set of variant categories for gene-based testing.

##### Variant categories for gene-based tests

1. **High confidence pLoF**: high-confidence LoF variants, as defined by LOFTEE ^137^ (LOFTEE HC).
2. **Damaging missense/protein-altering**: at least one of:

a. Variant annotated as missense/start-loss/stop-loss/in-frame indel and (REVEL≥0.773 or CADD≥28.1 (or both)).
b. Any variant with SpliceAI delta score (DS)≥0.2 where SpliceAI DS the maximum of the set {DS_AG, DS_AL, DS_DG, DS_DL} for each annotated variant (where DS_AG, DS_AL, DS_DG and DS_DL are delta score (acceptor gain), delta score (acceptor loss), delta score (donor gain), and delta score (donor loss), respectively).
c. Low-confidence LoF variants, as defined by LOFTEE (LOFTEE LC)
3. **Other missense/protein-altering**:

Missense/start-loss/stop-loss/in-frame indel not categorised in (2) (Damaging missense/protein-altering).
4. **Synonymous**: synonymous variants with SpliceAI DS<0.2 in the gene (our ‘control’ set).

REVEL and CADD score cut-offs are chosen to reflect the supporting level for pathogenicity (PP3) from the American College of Medical Genetics and Genomics and the Association for Molecular Pathology (ACMG/AMP) criteria^144^.

Variant counts and average allele counts for each annotation, split by population label and binned by MAF are displayed in Supp. Figure 13 and Supp. Figure 14, respectively.

#### Genetic association testing

We carried out rare variant genetic association testing in the European-ancestry subset of the UK Biobank using Scalable and Accurate Implementation of GEneralized mixed model (SAIGE) ^108^, a mixed model framework that accounts for sample relatedness and case-control imbalance through a saddle-point approximation in binary traits. All rare-variant analysis was carried out on the UK Biobank Research Analysis Platform (RAP) using SAIGE version wzhou88/saige:1.1.9^108^. In the sex-combined analyses, we account for *age, sex, age*^2^*, age × sex, age*^2^ *× sex*, and the first 10 genetic principal components as fixed effects; and *age, age*^2^, and the first 10 principal components in sex-specific analyses. All continuous traits were inverse rank normalised prior to association testing.

For SAIGE step 0, we constructed a genetic relatedness matrix (GRM) using the UK Biobank genotyping array data. We LD pruned the genotyped data using PLINK (`--indep-pairwise 50 5 0.05`)^145^, and created a sparse GRM using 5000 randomly selected markers, with relatedness cutoff of 0.05, using the `createSparseGRM.R` function within SAIGE. To generate a variance ratio file for subsequent steps in SAIGE, we extracted 1000 variants each with MAC<20 and MAC>20, and combined these markers to define a PLINK file for the variance ratio determination.

In SAIGE step 1 for each trait, the curated phenotype data and sparse GRM were used to fit a null model with no genetic contribution. All parameters were set at the defaults in SAIGE, except `--relatednessCutoff 0.05`, `--useSparseGRMtoFitNULL TRUE` and `--isCateVarianceRatio TRUE.` Tolerance for fitting the null generalised linear mixed model was set to 0.00001.

##### Rare variant and gene based testing

Following null model fitting, we carried out variant and gene-based testing in SAIGE step 2 using the variant categories described above, with the `--is_single_in_groupTest TRUE` flag. All other parameters were set to default, except `--maxMAF_in_groupTest=0.0001,0.001,0.01`, -`-is_Firth_beta TRUE`, `--pCutoffforFirth=0.1`, and -`-is_fastTest TRUE`. We included the following collection of group tests, using the annotations defined in methods: variant annotation.

- High confidence pLoF
- Damaging missense/protein-altering
- Other missense/protein-altering
- Synonymous
- High confidence pLoF or Damaging missense/protein-altering
- High confidence pLoF or Damaging missense/protein-altering or Other missense/protein-altering or Synonymous

We then carried out Cauchy combination tests^146^ across these annotations for each gene.

## Supporting information

FinnGen Banner Authors

## Data and code availability

Cohorts may be contacted individually for access to raw data. Summary statistics for all phenotypes will be made available through the GWAS Catalog upon publication. All code used in this study will be made available through GitHub upon publication.

## Acknowledgements

Thanks to Prof Pier Palamara and Prof Zoltan Kutalik for helpful discussions. We are grateful to the participants of all cohorts. This research has partly been conducted using the UK Biobank Resource under Application Number 11867. Genes & Health is/has recently been core-funded by Wellcome (WT102627, WT210561), the Medical Research Council (UK) (M009017, MR/X009777/1, MR/X009920/1), Higher Education Funding Council for England Catalyst, Barts Charity (845/1796), Health Data Research UK (for London substantive site), and research delivery support from the NHS National Institute for Health Research Clinical Research Network (North Thames). Genes & Health is/has recently been funded by Alnylam Pharmaceuticals, Genomics PLC; and a Life Sciences Industry Consortium of Astra Zeneca PLC, Bristol-Myers Squibb Company, GlaxoSmithKline Research and Development Limited, Maze Therapeutics Inc, Merck Sharp & Dohme LLC, Novo Nordisk A/S, Pfizer Inc, Takeda Development Centre Americas Inc. We thank Social Action for Health, Centre of The Cell, members of our Community Advisory Group, and staff who have recruited and collected data from volunteers. We thank the NIHR National Biosample Centre (UK Biocentre), the Social Genetic & Developmental Psychiatry Centre (King’s College London), Wellcome Sanger Institute, and Broad Institute for sample processing, genotyping, sequencing and variant annotation. We thank: Barts Health NHS Trust, NHS Clinical Commissioning Groups (City and Hackney, Waltham Forest, Tower Hamlets, Newham, Redbridge, Havering, Barking and Dagenham), East London NHS Foundation Trust, Bradford Teaching Hospitals NHS Foundation Trust, Public Health England (especially David Wyllie), Discovery Data Service/Endeavour Health Charitable Trust (especially David Stables), Voror Health Technologies Ltd (especially Sophie Don), NHS England (for what was NHS Digital) - for GDPR-compliant data sharing backed by individual written informed consent. Most of all we thank all of the volunteers participating in Genes & Health. This study was funded by the European Union through the European Regional Development Fund Project No. 2014-2020.4.01.15-0012 GENTRANSMED. Data analysis was carried out in part in the High-Performance Computing Center of University of Tartu. This research has partly been conducted using the ALSPAC resource under Project Number B4568. The activities of the EstBB are regulated by the Human Genes Research Act, which was adopted in 2000 specifically for the operations of the EstBB. Individual level data analysis in the EstBB was carried out under ethical approval 1.1-12/624 from the Estonian Committee on Bioethics and Human Research (Estonian Ministry of Social Affairs), using data according to release application 3-10/GI/10790 from the Estonian Biobank. This study was funded by the Estonian Research Council grants PRG1911. We want to acknowledge the participants and investigators of the FinnGen study. The FinnGen project is funded by two grants from Business Finland (HUS 4685/31/2016 and UH 4386/31/2016) and the following industry partners: AbbVie Inc., AstraZeneca UK Ltd, Biogen MA Inc., Bristol Myers Squibb (and Celgene Corporation & Celgene International II Sàrl), Genentech Inc., Merck Sharp & Dohme LCC, Pfizer Inc., GlaxoSmithKline Intellectual Property Development Ltd., Sanofi US Services Inc., Maze Therapeutics Inc., Janssen Biotech Inc, Novartis Pharma AG, and Boehringer Ingelheim International GmbH. Following biobanks are acknowledged for delivering biobank samples to FinnGen: Auria Biobank (www.auria.fi/biopankki), THL Biobank (www.thl.fi/biobank), Helsinki Biobank (www.helsinginbiopankki.fi), Biobank Borealis of Northern Finland (https://www.ppshp.fi/Tutkimus-ja-opetus/Biopankki/Pages/Biobank-Borealis-briefly-in-English.aspx), Finnish Clinical Biobank Tampere (www.tays.fi/en-US/Research_and_development/Finnish_Clinical_Biobank_Tampere), Biobank of Eastern Finland (www.ita-suomenbiopankki.fi/en), Central Finland Biobank (www.ksshp.fi/fi-FI/Potilaalle/Biopankki), Finnish Red Cross Blood Service Biobank (www.veripalvelu.fi/verenluovutus/biopankkitoiminta), Terveystalo Biobank (www.terveystalo.com/fi/Yritystietoa/Terveystalo-Biopankki/Biopankki/) and Arctic Biobank (https://www.oulu.fi/en/university/faculties-and-units/faculty-medicine/northern-finland-birth-cohorts-and-arctic-biobank). All Finnish Biobanks are members of BBMRI.fi infrastructure (www.bbmri.fi). Finnish Biobank Cooperative -FINBB (https://finbb.fi/) is the coordinator of BBMRI-ERIC operations in Finland. The Finnish biobank data can be accessed through the Fingenious® services (https://site.fingenious.fi/en/) managed by FINBB. We are extremely grateful to all the families who took part in this study, the midwives for their help in recruiting them, and the whole ALSPAC team, which includes interviewers, computer and laboratory technicians, clerical workers, research scientists, volunteers, managers, receptionists and nurses. We acknowledge the contribution of the participants in the deCODE study. Patients and control subjects in FinnGen provided informed consent for biobank research, based on the Finnish Biobank Act. Alternatively, separate research cohorts, collected prior the Finnish Biobank Act came into effect (in September 2013) and start of FinnGen (August 2017), were collected based on study-specific consents and later transferred to the Finnish biobanks after approval by Fimea (Finnish Medicines Agency), the National Supervisory Authority for Welfare and Health. Recruitment protocols followed the biobank protocols approved by Fimea. The Coordinating Ethics Committee of the Hospital District of Helsinki and Uusimaa (HUS) statement number for the FinnGen study is Nr HUS/990/2017. The FinnGen study is approved by Finnish Institute for Health and Welfare (permit numbers: THL/2031/6.02.00/2017, THL/1101/5.05.00/2017, THL/341/6.02.00/2018, THL/2222/6.02.00/2018, THL/283/6.02.00/2019, THL/1721/5.05.00/2019 and THL/1524/5.05.00/2020), Digital and population data service agency (permit numbers: VRK43431/2017-3, VRK/6909/2018-3, VRK/4415/2019-3), the Social Insurance Institution (permit numbers: KELA 58/522/2017, KELA 131/522/2018, KELA 70/522/2019, KELA 98/522/2019, KELA 134/522/2019, KELA 138/522/2019, KELA 2/522/2020, KELA 16/522/2020), Findata permit numbers THL/2364/14.02/2020, THL/4055/14.06.00/2020, THL/3433/14.06.00/2020, THL/4432/14.06/2020, THL/5189/14.06/2020, THL/5894/14.06.00/2020, THL/6619/14.06.00/2020, THL/209/14.06.00/2021, THL/688/14.06.00/2021, THL/1284/14.06.00/2021, THL/1965/14.06.00/2021, THL/5546/14.02.00/2020, THL/2658/14.06.00/2021, THL/4235/14.06.00/2021, Statistics Finland (permit numbers: TK-53-1041-17 and TK/143/07.03.00/2020 (earlier TK-53-90-20) TK/1735/07.03.00/2021, TK/3112/07.03.00/2021) and Finnish Registry for Kidney Diseases permission/extract from the meeting minutes on 4th July 2019. The Biobank Access Decisions for FinnGen samples and data utilised in FinnGen Data Freeze 10 include: THL Biobank BB2017_55, BB2017_111, BB2018_19, BB_2018_34, BB_2018_67, BB2018_71, BB2019_7, BB2019_8, BB2019_26, BB2020_1, BB2021_65, Finnish Red Cross Blood Service Biobank 7.12.2017, Helsinki Biobank HUS/359/2017, HUS/248/2020, HUS/150/2022 § 12, §13, §14, §15, §16, §17, §18, and §23, Auria Biobank AB17-5154 and amendment #1 (August 17 2020) and amendments BB_2021-0140, BB_2021-0156 (August 26 2021, Feb 2 2022), BB_2021-0169, BB_2021-0179, BB_2021-0161, AB20-5926 and amendment #1 (April 23 2020)and it’s modification (Sep 22 2021), Biobank Borealis of Northern Finland_2017_1013, 2021_5010, 2021_5018, 2021_5015, 2021_5023, 2021_5017, 2022_6001, Biobank of Eastern Finland 1186/2018 and amendment 22 § /2020, 53§/2021, 13§/2022, 14§/2022, 15§/2022, Finnish Clinical Biobank Tampere MH0004 and amendments (21.02.2020 & 06.10.2020), §8/2021, §9/2022, §10/2022, §12/2022, §20/2022, §21/2022, §22/2022, §23/2022, Central Finland Biobank 1-2017, and Terveystalo Biobank STB 2018001 and amendment 25th Aug 2020, Finnish Hematological Registry and Clinical Biobank decision 18th June 2021, Arctic biobank P0844: ARC_2021_1001. We want to acknowledge the participants and investigators of FinnGen study. The FinnGen project is funded by two grants from Business Finland (HUS 4685/31/2016 and UH 4386/31/2016) and the following industry partners: AbbVie Inc., AstraZeneca UK Ltd, Biogen MA Inc., Bristol Myers Squibb (and Celgene Corporation & Celgene International II Sàrl), Genentech Inc., Merck Sharp & Dohme LCC, Pfizer Inc., GlaxoSmithKline Intellectual Property Development Ltd., Sanofi US Services Inc., Maze Therapeutics Inc., Janssen Biotech Inc, Novartis Pharma AG, and Boehringer Ingelheim International GmbH. Following biobanks are acknowledged for delivering biobank samples to FinnGen: Auria Biobank (www.auria.fi/biopankki), THL Biobank (www.thl.fi/biobank), Helsinki Biobank (www.helsinginbiopankki.fi), Biobank Borealis of Northern Finland (https://www.ppshp.fi/Tutkimus-ja-opetus/Biopankki/Pages/Biobank-Borealis-briefly-in-English.aspx), Finnish Clinical Biobank Tampere (www.tays.fi/en-US/Research_and_development/Finnish_Clinical_Biobank_Tampere), Biobank of Eastern Finland (www.ita-suomenbiopankki.fi/en), Central Finland Biobank (www.ksshp.fi/fi-FI/Potilaalle/Biopankki), Finnish Red Cross Blood Service Biobank (www.veripalvelu.fi/verenluovutus/biopankkitoiminta), Terveystalo Biobank (www.terveystalo.com/fi/Yritystietoa/Terveystalo-Biopankki/Biopankki/) and Arctic Biobank (https://www.oulu.fi/en/university/faculties-and-units/faculty-medicine/northern-finland-birth-cohorts-and-arctic-biobank). All Finnish Biobanks are members of BBMRI.fi infrastructure (www.bbmri.fi). Finnish Biobank Cooperative -FINBB (https://finbb.fi/) is the coordinator of BBMRI-ERIC operations in Finland. The Finnish biobank data can be accessed through the Fingenious® services (https://site.fingenious.fi/en/) managed by FINBB. This study was based on the CHB reproduction protocol and DBDS (ethical approval NVK-1805807; NVK-1700407; SJ-740).

## Supplementary Author List

Genes & Health Research Team: Shaheen Akhtar, Mohammad Anwar, Elena Arciero, Omar Asgar, Samina Ashraf, Saeed Bidi, Gerome Breen, James Broster, Raymond Chung, David Collier, Charles J Curtis, Shabana Chaudhary, Megan Clinch, Grainne Colligan, Panos Deloukas, Ceri Durham, Faiza Durrani, Fabiola Eto, Sarah Finer, Joseph Gafton, Ana Angel, Chris Griffiths, Joanne Harvey, Teng Heng, Sam Hodgson, Qin Qin Huang, Matt Hurles, Karen A Hunt, Shapna Hussain, Kamrul Islam, Vivek Iyer, Benjamin M Jacobs, Ahsan Khan, Claudia Langenberg, Cath Lavery, Sang Hyuck Lee, Daniel MacArthur, Sidra Malik, Daniel Malawsky, Hilary Martin, Dan Mason, Rohini Mathur, Mohammed Bodrul Mazid, John McDermott, Caroline Morton, Bill Newman, Elizabeth Owor, Asma Qureshi, Shwetha Ramachandrappa, Mehru Raza, Jessry Russell, Nishat Safa, Miriam Samuel, Michael Simpson, John Solly, Marie Spreckley. Daniel Stow, Michael Taylor, Richard C Trembath, Karen Tricker, David A van Heel, Klaudia Walter, Caroline Winckley, Suzanne Wood, John Wright, Ishevanhu Zengeya, Julia Zöllner.

Estonian Biobank Research Team: Andres Metspalu, Lili Milani, Tõnu Esko, Mari Nelis and Georgi Hudjashov.

Estonian Health Information Research Team: Raivo Kolde, Sven Laur, Sulev Reisberg and Jaak Vilo.

DBDS Genomic Consortium: Agnete Lundgaard; Alexander Pil Henriksen; Bertram Dalskov Kjerulff; Bitten Aagaard Jensen; Bjarke Feenstra; Christian Erikstrup; Christina Mikkelsen; Daniel Gudbjartsson; David Westergaard; Erik Sørensen; Frank Geller; Gregor Jemec; Henrik Hjalgrim; Henrik Ullum; Hreinn Stefánsson; Ioanna Nissen; Ioannis Louloudis ; Jakob Bay; Jens Kjærgaard Boldsen; Joseph Dowsett; Kari Stefansson; Karina Banasik; Katrine Kaspersen; Khoa Manh Dinh; Klaus Rostgaard; Kristoffer Burgdorf; Lise Wegner Thørner; Lisette Kogelman; Lotte Hindhede; Margit Anita Hørup Larsen; Maria Didriksen; Mette Nyegaard; Michael Schwinn; Mie Topholm Bruun; Mona Ameri Chalmer; Ole Birger Pedersen; Palle Duun Rohde; Rikke Louise Jacobsen; Sisse Rye Ostrowski; Søren Brunak; Susan Mikkelsen; Thomas Folkmann Hansen; Thomas Werge; Thorsten Brodersen; Unnur Þorsteinsdóttir.

FinnGen: See separate document.

