## Supplemental Figures for "Genome-wide analyses identify 21 infertility loci and over 400 reproductive hormone loci across the allele frequency spectrum"

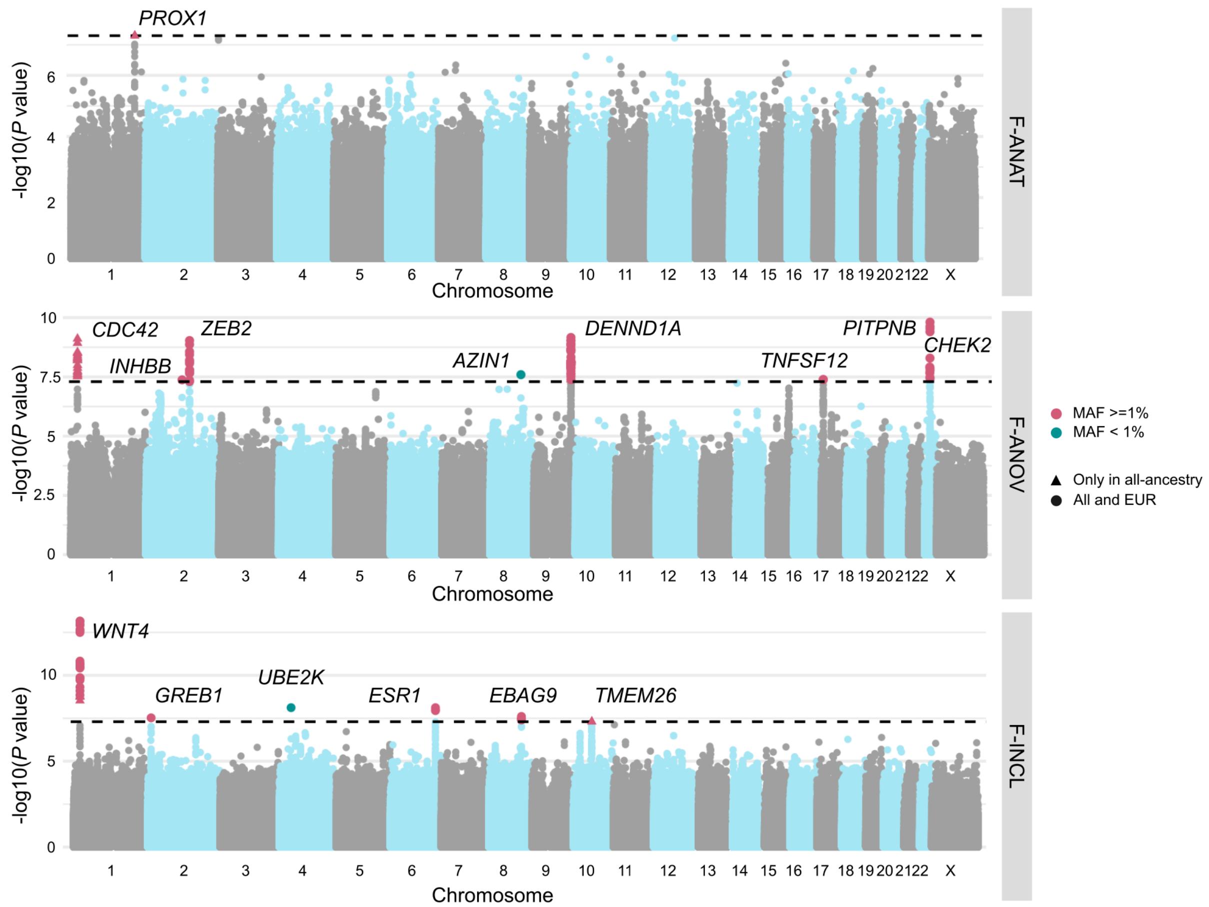


**Supp. Figure 1. Manhattan plots for female infertility GWAS meta-analyses not presented in the main text.** Genetic variants associated with anatomical female infertility (F-ANAT) (top), anovulatory female infertility (F-ANOV) (middle), and idiopathic infertility (unknown causes) defined by inclusion of a code for idiopathic infertility (F-INCL) (bottom). Each point depicts a single SNP, with genome-wide significant (GWS) SNPs (P < 5E-08, dashed line) coloured in pink for common variants with minor allele frequency (MAF)>=1% and green for those with MAF<1%. Triangles represent SNPs that only reach GWS in all-ancestry GWAS meta-analyses. SNPs are annotated with the mapped gene.

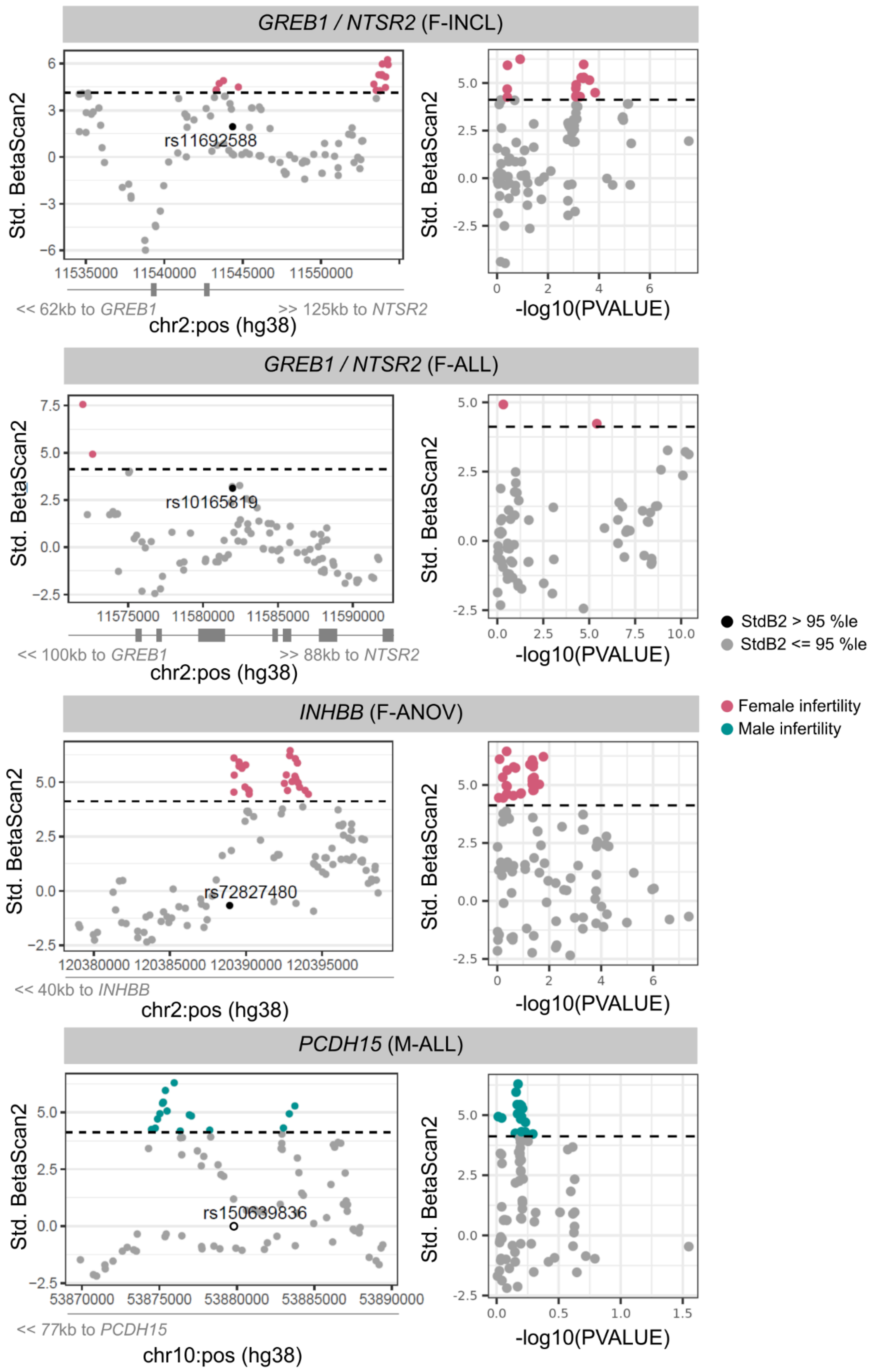


**Supp. Figure 2. Balancing selection, as measured by standardised BetaScan2 (StdB2) scores, at infertility-associated loci.** Each panel displays windows of +/- 10 kb around a lead infertility-associated variant, annotated with nearest gene and location: rs10165819 (F-ALL), rs72827480 (anovulatory infertility, F-ANOV), rs11692588 (female idiopathic infertility by inclusion, F-INCL), and rs150639836 (male infertility of all causes, M-ALL). Dashed lines indicate 95th %ile of

StdB2, and variants crossing this threshold are coloured in pink (for female infertility loci) or green (for the male infertility locus). Left: Locus plots depicting genomic position on the x-axis and StdB2 on the y-axis. The lead variant rs150639836 (open circle) is not present in the StdB2 dataset and thus assigned StdB2 of 0. Right: Scatter plots depicting relationship between -log10 of the GWAS p-value for the variant association with infertility on the x-axis and StdB2 on the y-axis.

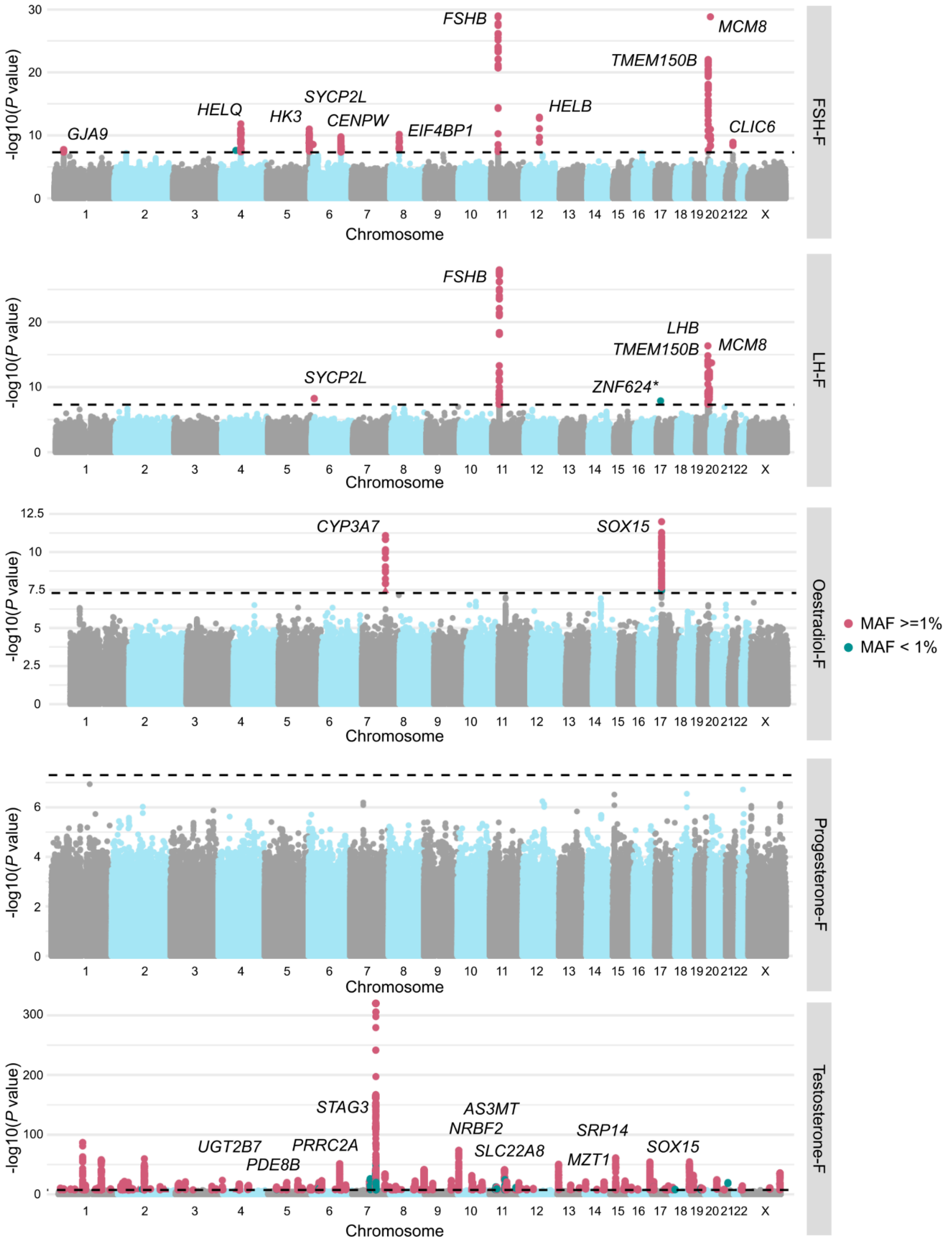


**Supp. Figure 3. Manhattan plots for female reproductive hormone GWAS meta-analyses.** Each panel displays genetic variants associated with a different reproductive hormone, from top to bottom: follicle stimulating hormone (FSH), luteinising hormone (LH), oestradiol, progesterone, and total testosterone. Each point depicts a single SNP, with genome-wide significant (GWS) SNPs (P < 5E-08, dashed line) coloured: in pink for common variants with minor allele frequency (MAF)>=1% and green for those with MAF<1%. SNPs are annotated with the mapped gene; for testosterone, only novel SNPs (ten lowest P-values) are annotated. * indicates that lead variant is present in only one study. For all lead variants, refer to Supp. Table 10.

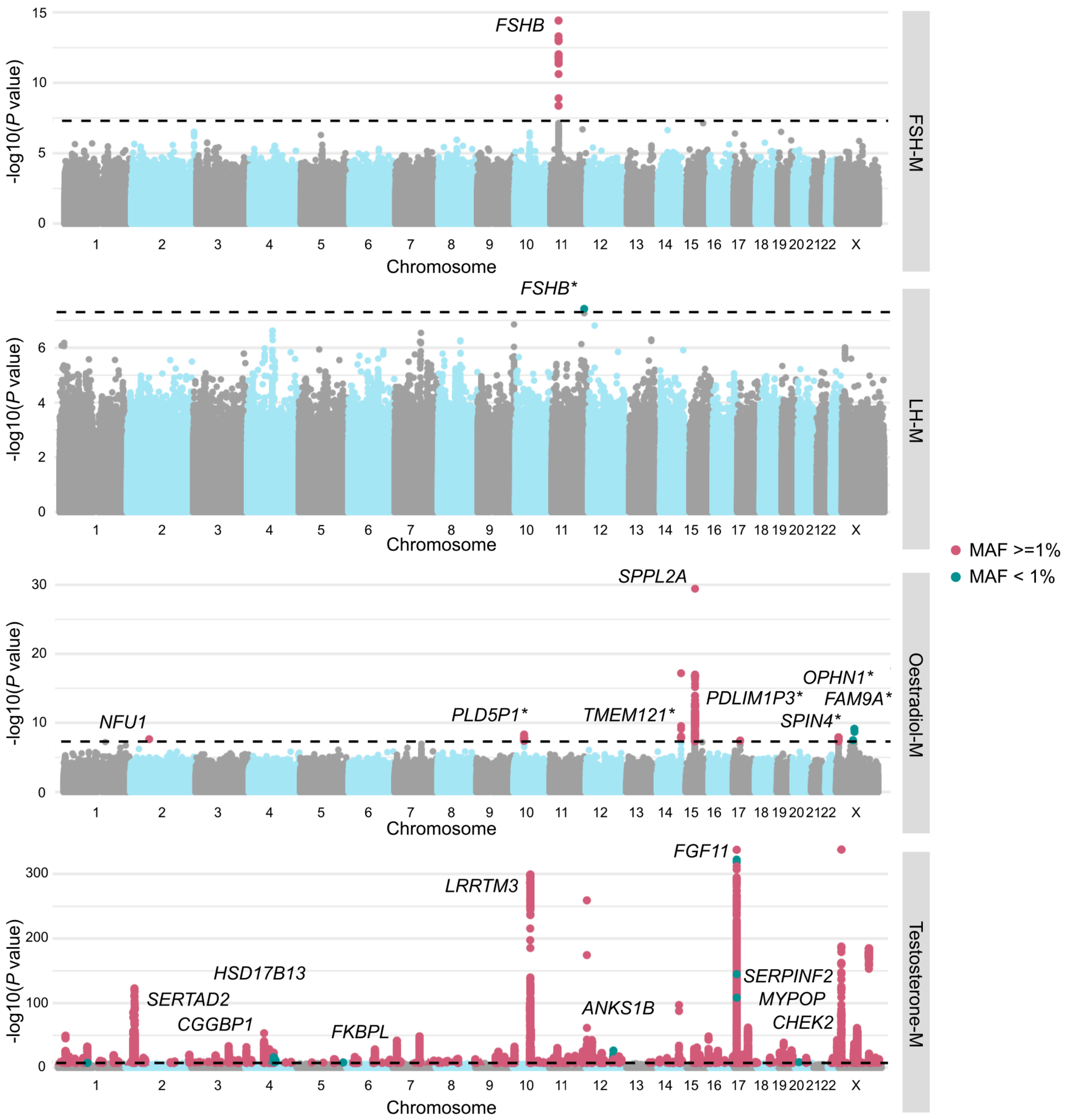


**Supp. Figure 4. Manhattan plots for male reproductive hormone GWAS meta-analyses.** Each panel displays genetic variants associated with a different reproductive hormone, from top to bottom: follicle stimulating hormone (FSH), luteinising hormone (LH), oestradiol, and total testosterone. Each point depicts a single SNP, with genome-wide significant (GWS) SNPs (P < 5E-08, dashed line) coloured: in pink for common variants with minor allele frequency (MAF)>=1% and green for those with MAF<1%. SNPs are annotated with the mapped gene; for testosterone, only novel SNPs (ten lowest P-values) are annotated. * indicates that lead variant is present in only one study. For all lead variants, refer to Supp. Table 10.

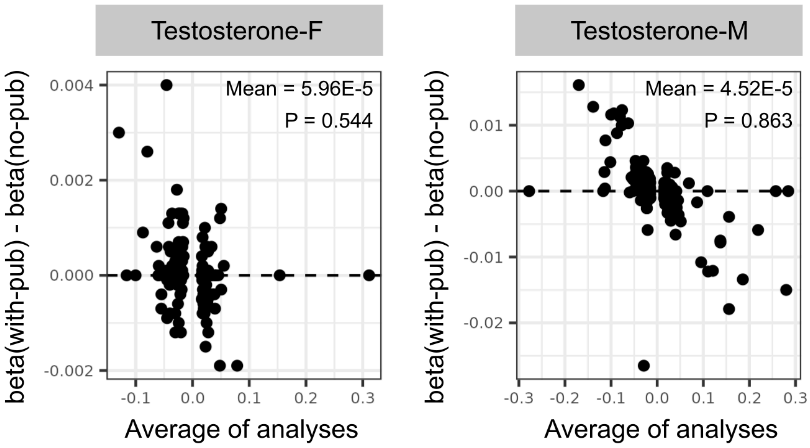


**Supp. Figure 5. Comparison of effect sizes of SNPs associated with testosterone in main meta-analyses and sensitivity analyses without publicly available summary statistics.** Variants that are genome-wide significant (GWS *P* < 5E-08) in the main GWAS with public data are plotted. The Bland-Altman plot displays the difference between effect sizes estimated in the with-public data (with-pub) and no-public data (no-pub) GWASs for each variant, plotted against the mean estimate from the two sets of analyses. The mean difference and one-sample t-test P-value (with null hypothesis of mean difference = 0) are displayed for each of the strata. Left: female-specific analyses, Right: male-specific analyses, from all-ancestry samples.

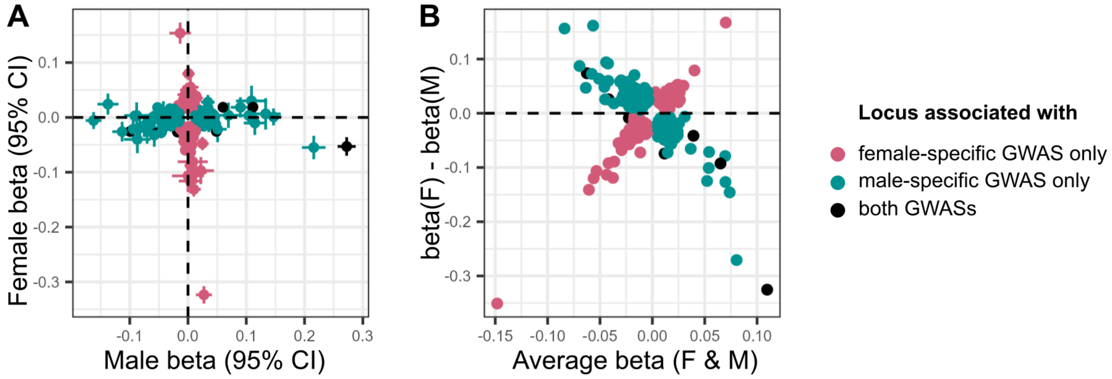


**Supp. Figure 6. Sex heterogeneity in the effects of lead variants associated with testosterone.** Lead variants in either female-specific testosterone GWAS (coloured in pink), male-specific testosterone GWAS (coloured in green), or both (coloured in black) are displayed. Results are from all-ancestry meta-analyses. (A) Scatter plot comparing effect sizes and 95% confidence intervals (CIs), plotted as error bars, of GWS variants. (B) Bland-Altman plot displaying the difference between effect sizes estimated in the female-specific and male-specific GWASs for each variant, plotted against the mean estimate from both GWASs.

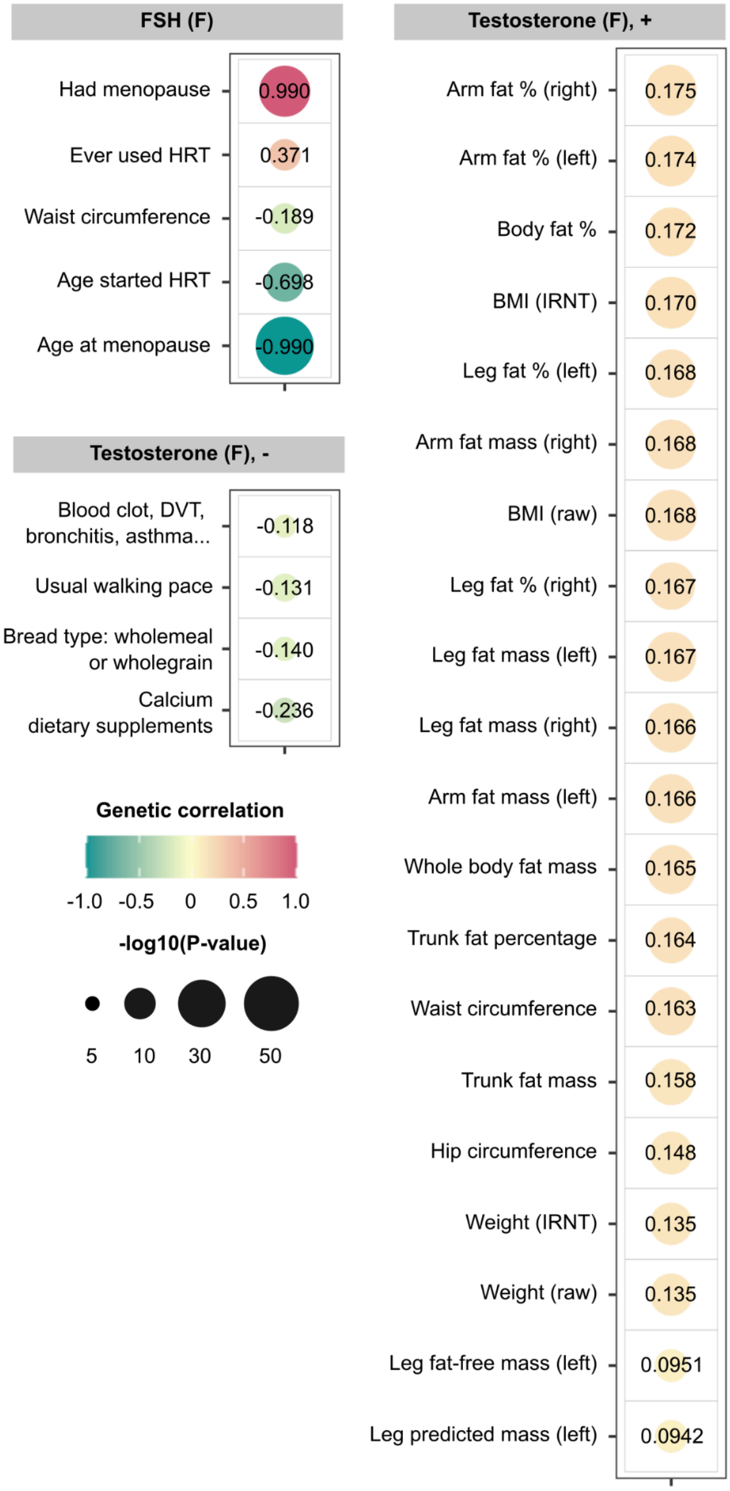


**Supp. Figure 7. Genetic correlations ($r_g$) between follicle stimulating hormone in females (FSH-F) or testosterone-F and traits across the phenome.** Trait summary statistics were generated by the Neale lab[53] and SNP-based genetic correlation calculations were performed using the LDSC software on a subset of 1 million HapMap3 SNPs[52]. Points are coloured by $r_g$ estimate and sized by -log10($P$). Only phenotypes associated at $P$ < 2.45E-05 (FWER controlled at 5% across 2,040 tests using the Bonferroni method, accounting for 340 effectively independent UKBB phenotypes and 6 hormone or infertility strata are displayed, up to a maximum of 20 phenotypes. Phenotypes in the panel labelled “Testosterone (F), -” are negatively correlated with testosterone-F, and those in the panel labelled “Testosterone (F), +” are positively correlated with testosterone-F. HRT = hormone replacement therapy, BMI = body mass index, IRNT = inverse-rank normally transformed, “Blood clot, DVT, bronchitis, asthma…” = Blood clot, deep vein thrombosis (DVT), bronchitis, emphysema, asthma, rhinitis, eczema, Hayfever, allergic rhinitis or eczema allergy diagnosed by doctor.

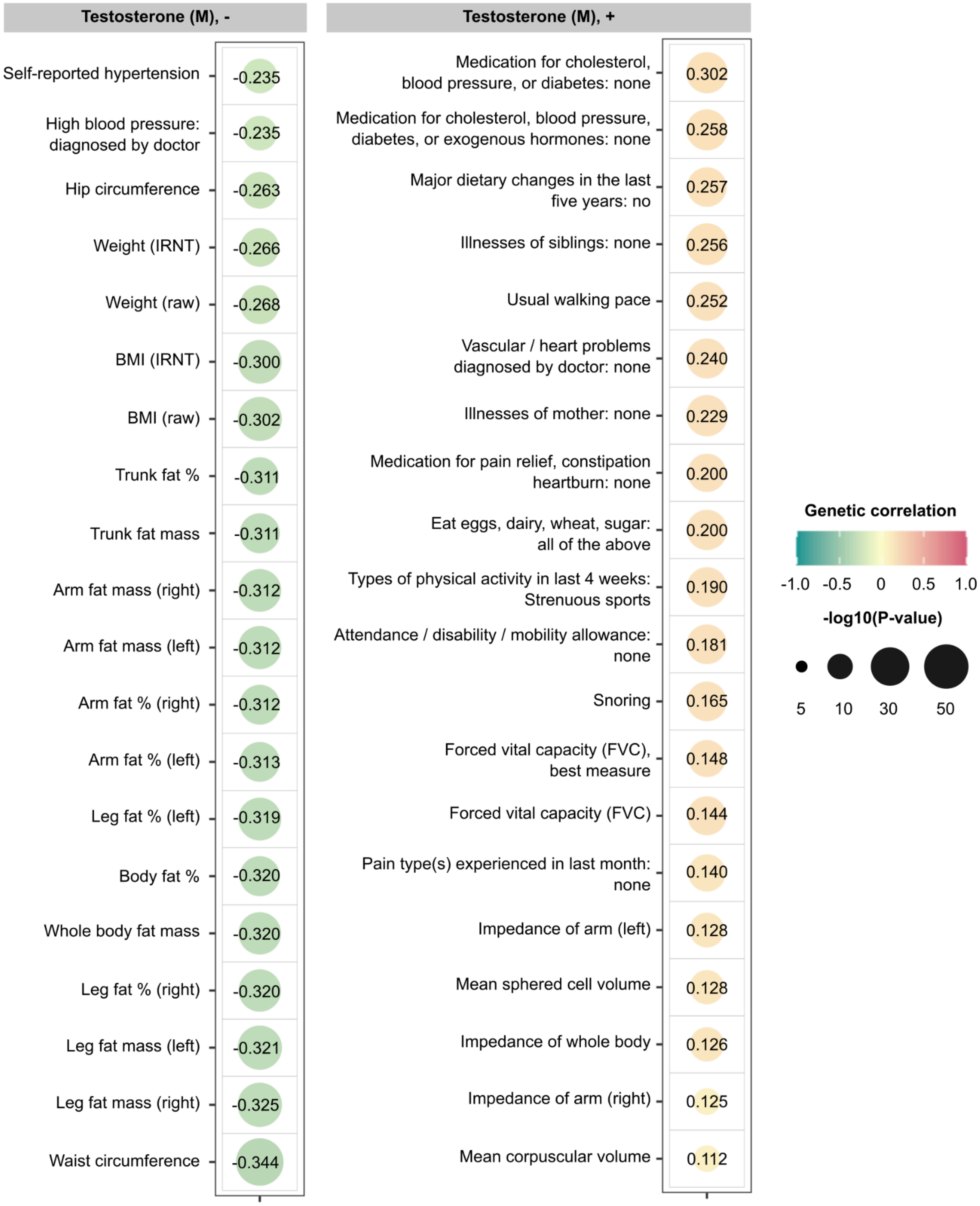


**Supp. Figure 8. Genetic correlations ($r_g$) between testosterone in males and traits across the phenome.** Trait summary statistics were generated by the Neale lab[53] and SNP-based genetic correlation calculations were performed using the LDSC software[51] on a subset of 1 million HapMap3 SNPs[52]. Points are coloured by $r_g$ estimate and sized by -log10($P$). Only phenotypes associated at $P$

< 2.45E-05 (FWER controlled at 5% across 2,040 tests using the Bonferroni method, accounting for 340 effectively independent UKBB phenotypes and 6 hormone or infertility strata are displayed, up to a maximum of 20 phenotypes. Phenotypes in the panel labelled “Testosterone (M), -” are negatively correlated with testosterone-M, and those in the panel labelled “Testosterone (M), +” are positively correlated with testosterone-M. BMI = body mass index, IRNT = inverse-rank normally transformed.

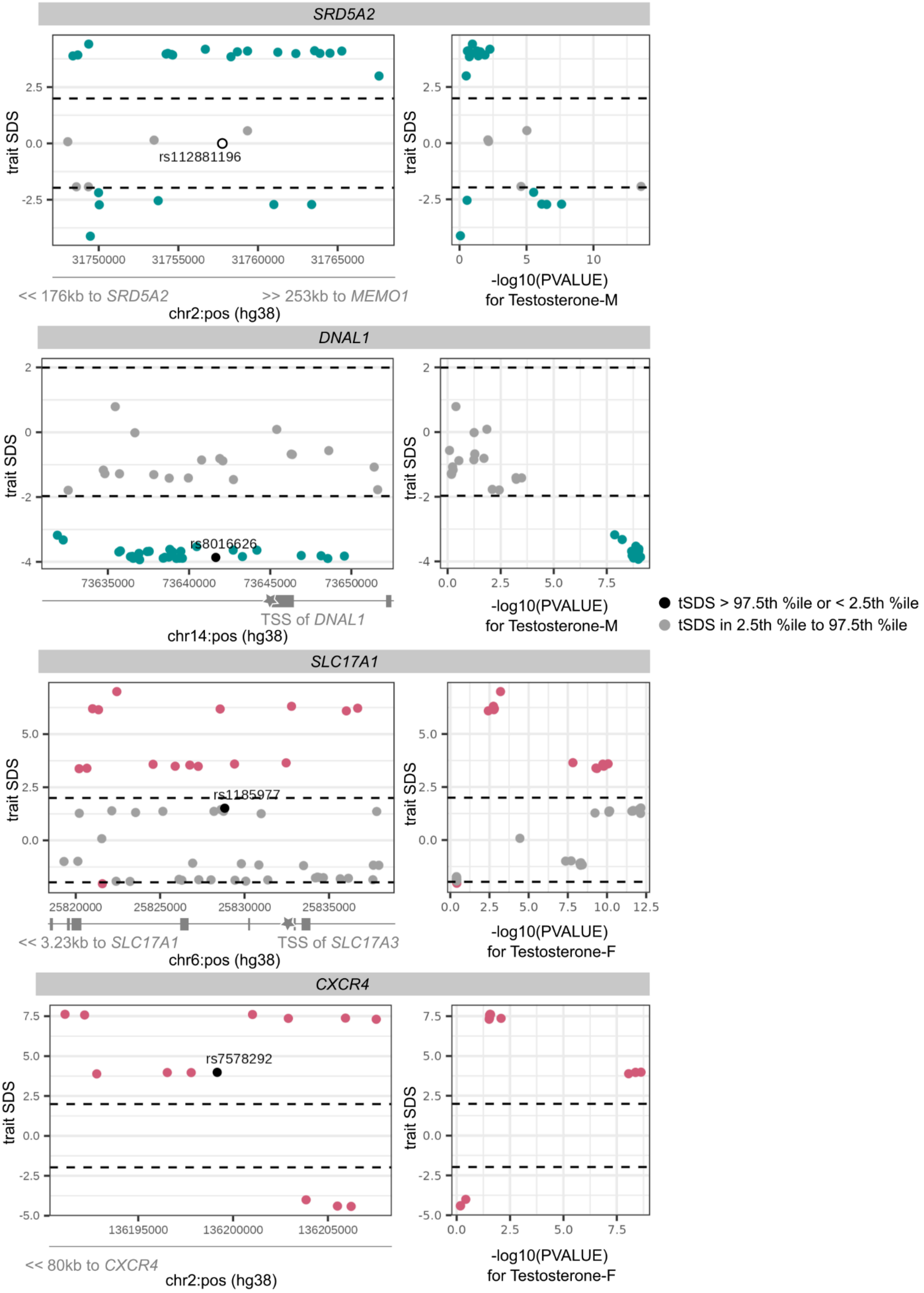


**Supp. Figure 9. Trait-aligned Singleton Density Scores (SDSs), measuring recent directional selection, at testosterone-associated loci.** Each panel displays windows of +/- 10 kb around a lead testosterone-associated variant, annotated with the location of nearest gene transcription start sites (TSSs) for all variants with extreme tSDSs: rs112881196 and rs8016626 (male-specific), and rs1185977 and rs7578292 (female-specific). The tSDSs are aligned to the testosterone-increasing allele, wherein a positive tSDS indicates positive selection for testosterone-increasing allele at the locus. Dashed lines indicate 2.5th percentile (%ile) and 97.5th %ile of SDSs, and variants below or above this threshold respectively are coloured in green (for male-specific loci) and pink (for female-specific loci). Left: Locus plots depicting genomic position on the x-axis and trait-SDS on the y-axis. The lead variant rs112881196 (open circle) is not present in the tSDS dataset and thus assigned a score of 0. Right: Scatter plots depicting relationship between -log10 of the GWAS p-value for the variant association with testosterone on the x-axis and tSDS on the y-axis.

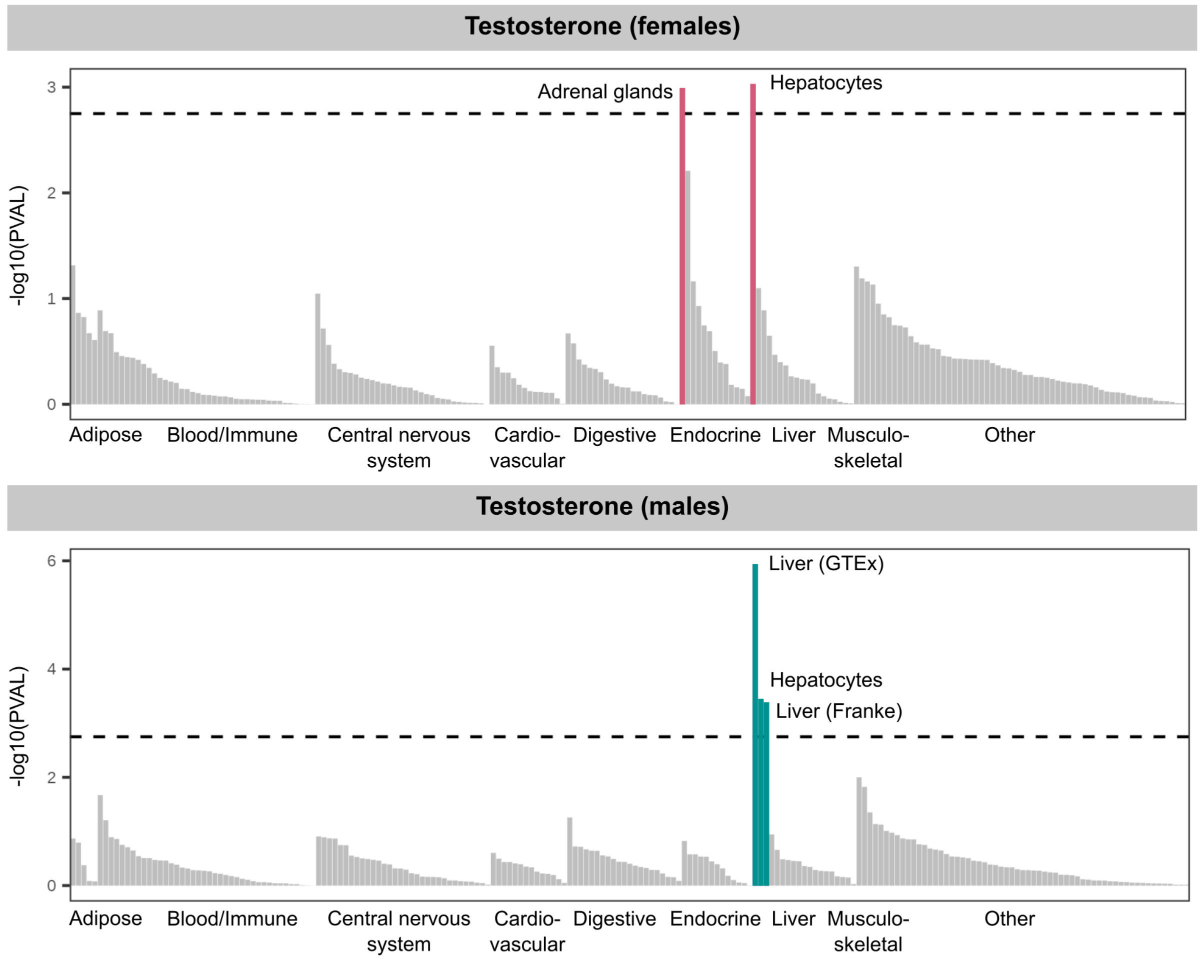


**Supp. Figure 10. Enrichment of testosterone heritability across tissues and cell types.** Partitioned heritability across 205 tissues and cell-types from the Genotype Tissue Expression (GTEx) Project database[41] and the Franke lab single-cell database[72] was assessed using partitioned LD-score regression[51]. Tissues and cell types are broadly grouped by organ system, and those that reach significance (FDR < 5%, dashed line), are annotated and coloured in: pink for testosterone in females (top) and green for testosterone in males (bottom).

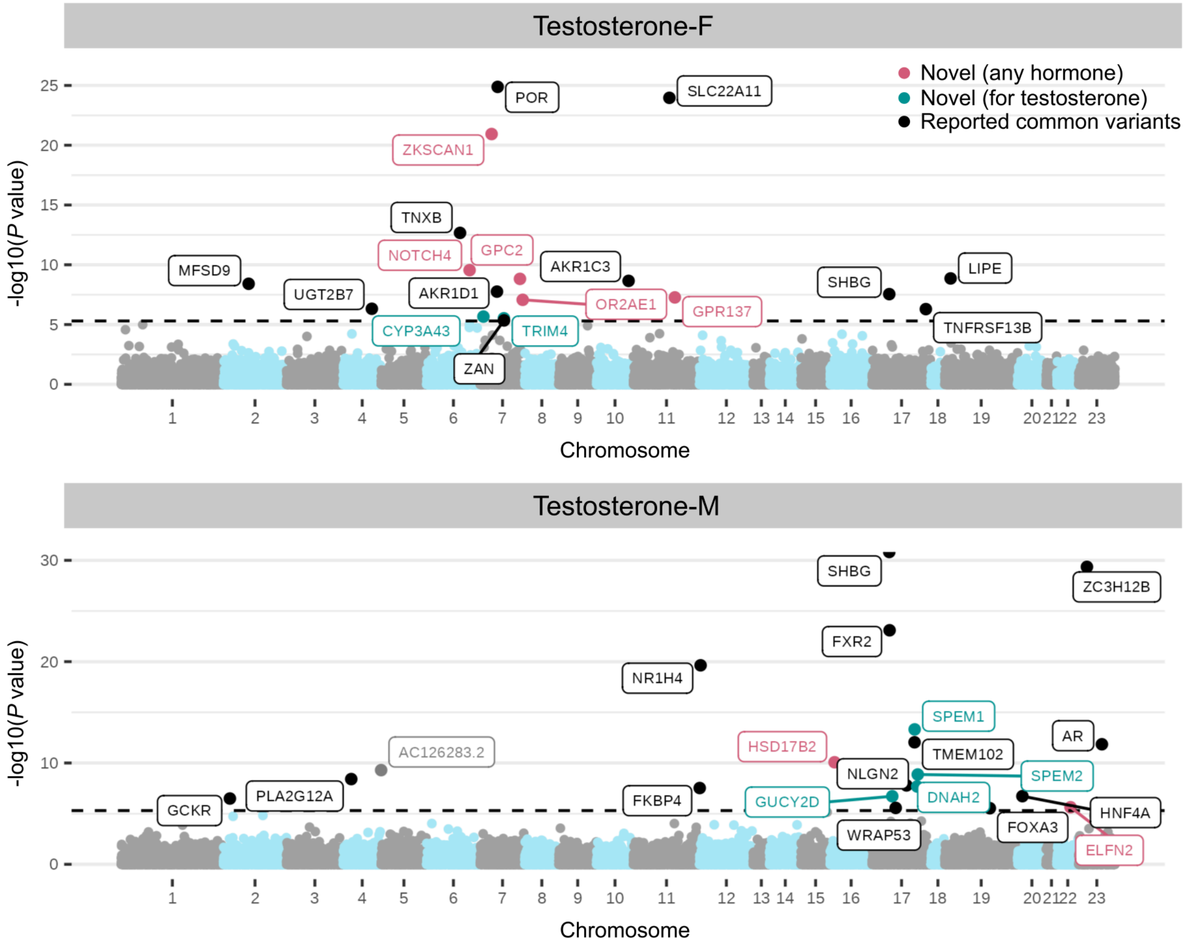


**Supp. Figure 11. Gene-based Manhattan plots for burden of rare variants associated with testosterone in females and males in UK Biobank.** Significance levels estimated using the Cauchy combined P-value are displayed, with significant genes (exome-wide significant at $P < 5E\text{-}06$ (FWER controlled at 5% using the Bonferroni method, across 10,000 effectively independent genes) coloured in: pink if no previous common variant associations with the gene have been reported for any of 28 reproductive hormones in the GWAS Catalog[62], green if no previous common variant associations with the gene have been reported for testosterone, and black otherwise.

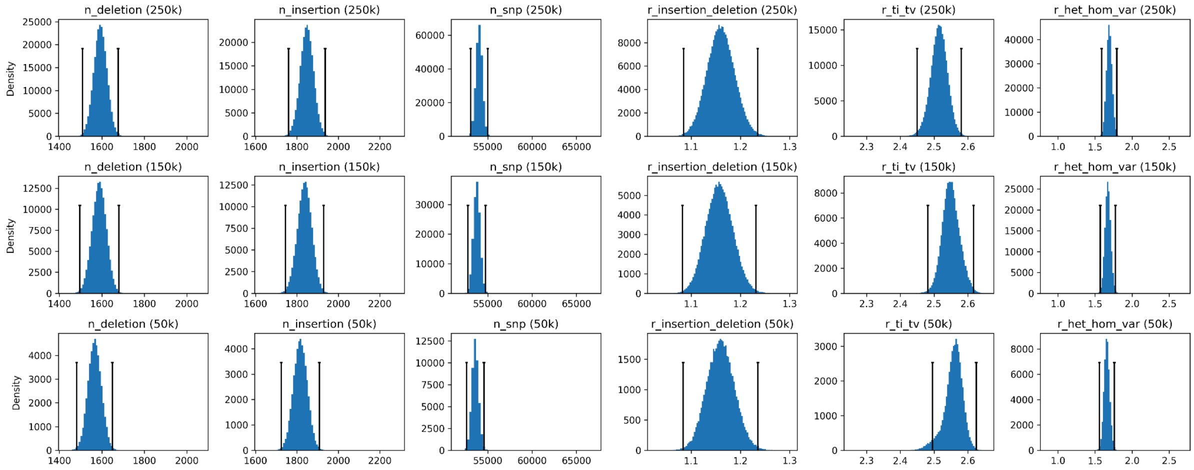


**Supp. Figure 12. MAD thresholds for QC of exome sequencing samples.** Samples with any of n_deletion, n_insertion, n_snp, r_insertion_deletion, r_ti_tv, and r_het_hom_var exceeding four MADs from the median are removed. MAD thresholds are displayed as vertical lines, conditional on tranche size (50k, 200k, 450k), from shortest to tallest.

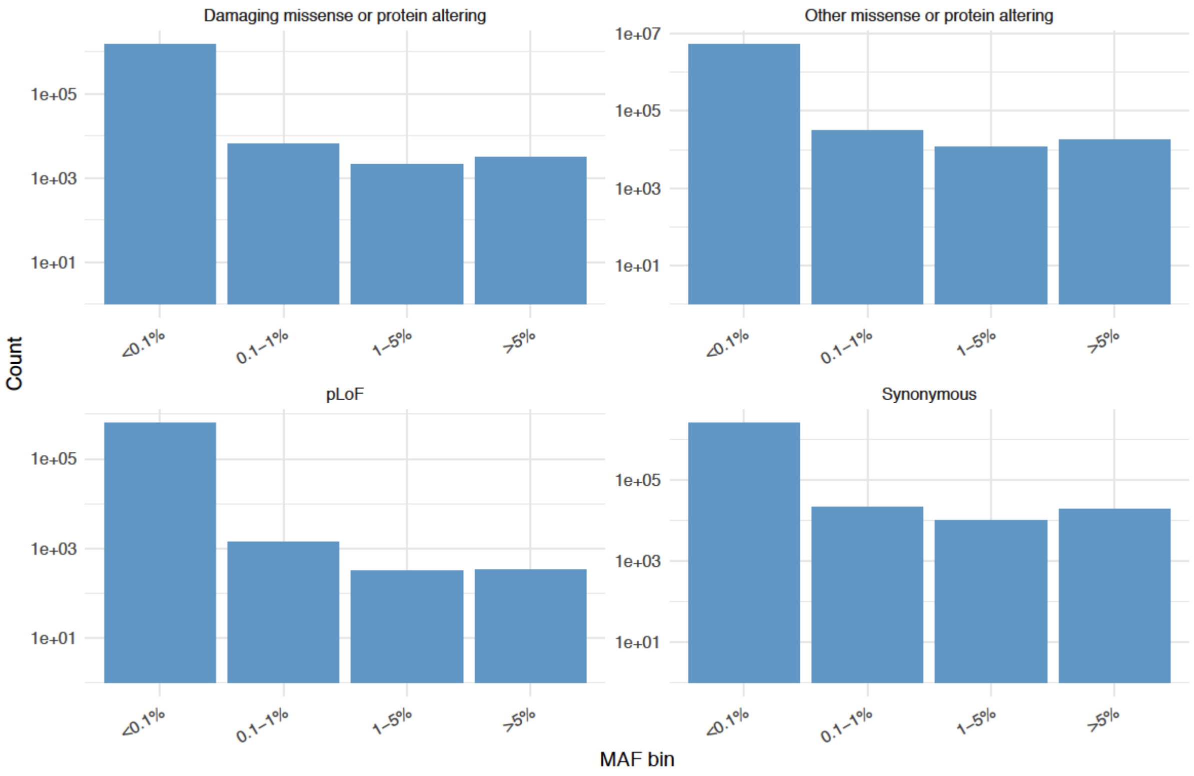


**Supp. Figure 13. Average number of variants per individual, binned by MAF, according to the x-axes.** Counts (y-axes) are on a log scale.

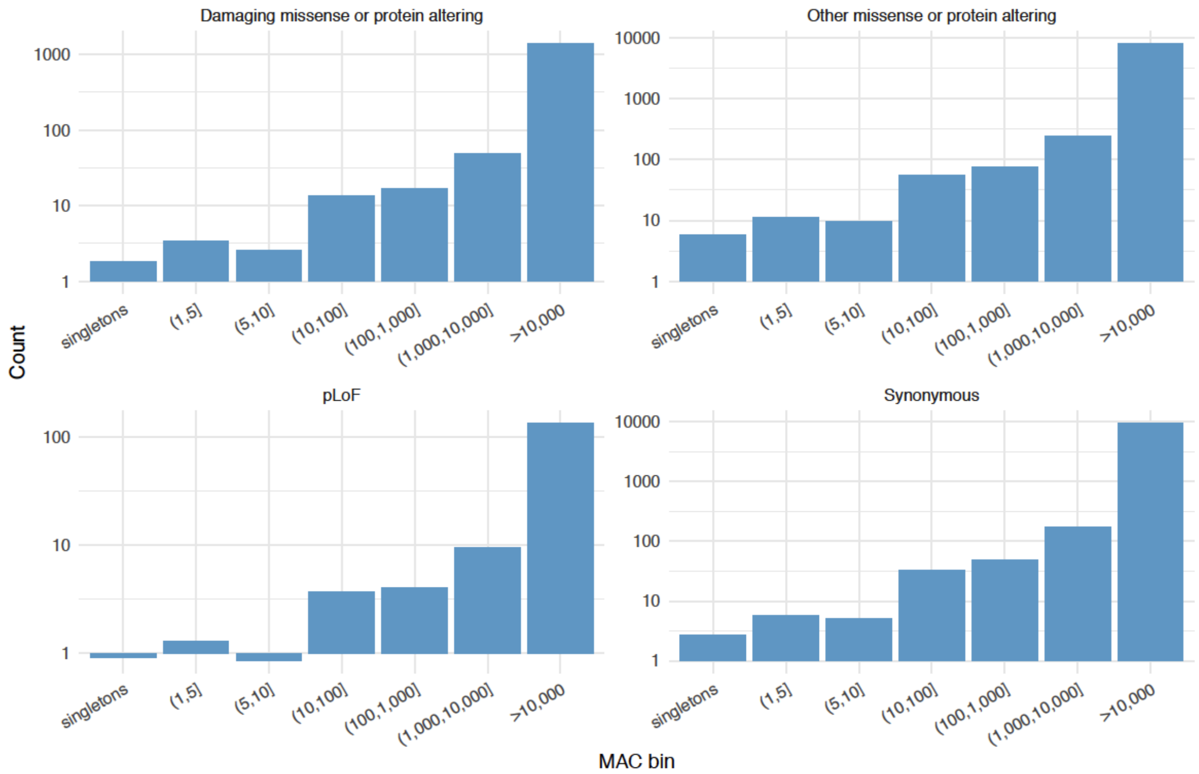


**Supp. Figure 14. Average number of variants binned by minor allele count (MAC), per individual according to the *x*-axes.** Count (y-axes) are on a log scale.
