## Supplemental Text for "Genome-wide analyses identify 21 infertility loci and over 400 reproductive hormone loci across the allele frequency spectrum"

### Supplementary Results

#### No evidence for heterogeneity in variant effects on infertility or reproductive hormones across cohorts

All reported lead variants for infertility were present in at least two cohorts (Supp. Table 3). Among these were three variants for F-ANOV, two variants for F-INCL, and one variant for M-ALL that were only present in FinnGen and EstBB; as the Finnish population underwent a recent bottleneck event and is genetically connected to Estonians, common variants in these two populations may not be represented in other European-ancestry cohorts[147]. Eighteen of 21 lead variants show no evidence for heterogeneity in effect estimates across cohorts (all $P_{het} > 0.05$), and the remaining have consistent effect directions in all of the studies in which they are present (Supp. Table 3).

We ensured that our hormone GWAS meta-analysis results were robust to the inclusion of summary statistics from publicly available datasets. The majority of lead variants associated with Oestradiol-F (2/2), Oestradiol-M (2/4), Testosterone-F (146/153), and Testosterone-M (230/243), were recovered at genome wide significance when publicly available datasets were excluded, and the small number of remaining variants were still nominally associated at $P$<5E-05. There is also no heterogeneity between effect size estimates from meta-analyses with and without public summary statistics (heterogeneity $P_{\text{het}}$>0.01 at all lead variants across all strata) (Supp. Figure 5). Finally, as with the infertility meta-analyses, all reported lead variants from the reproductive hormone meta-analyses were present in at least two of the cohorts examined and the majority (81%) show no evidence for heterogeneity in effect estimates across cohorts ($P_{\text{het}}$>0.05).

#### No evidence for genome-wide directional selection against infertility

We did not observe any GWS variants for infertility in 54 genomic regions under historical directional selection (50,000 years or 10,000 years) (Supp. Table 8). We also did not find significant genome-wide directional selection against infertility when SDS were aligned to the infertility-risk increasing allele: the mean genome-wide trait-SDS is not significantly different from 0 for any category of infertility (all $P$>0.05).

#### Variants associated with reproductive hormones in hormone metabolism genes

We replicated genetic variants associated with reproductive hormones in or near genes that encode the hormone subunits themselves, such as *FSHB,* which encodes the FSH beta subunit (rs11031005, FSH-F β (SE)=-0.104 (0.0092), $P$=1.08E-29 and FSH-M β=-0.206 (0.0263), $P$=3.75E-15), and *LHB*, which encodes the LH beta subunit (rs753307, LH-F β (SE)=0.0605 (0.0072), $P$=4.53E-17) (Supp. Table 10). We also replicated associations near genes encoding enzymes for steroid hormone metabolism, such as *CYP3A7*, whose encoded enzyme metabolises a precursor of oestrogen (rs45446698, Oestradiol-F β=-0.0782 (0.0114), $P$=8.26E-12), and *HSD17B13*, whose encoded protein catalyses the oxidation of oestradiol[148] (rs13133311, Testosterone-M β=-0.0307 (0.00270), $P$=2.84E-29) (Supp. Table 10).

#### Sex-specific genetic architecture of testosterone

Over a third (39.7%) of lead variants identified in either of the sex-specific analyses for testosterone had significant sex-differential effects (sex heterogeneity *P*<0.05/399, Bonferroni-adjusted for number of unique lead variants tested) (Supp. Figure 6). The genome-wide SNP-based heritability of total testosterone was 1.5x higher in men (15.67%, SE=1.79%) than in women (9.87% (1.00%)) (Supp. Table 6). The phenome-wide genetic correlation landscape of testosterone, assessed across 703 heritable phenotypes in UKBB, was also substantially different between the sexes (Supp. Figure 7 and Supp. Figure 8, phenome-wide significant at *P*<4.90E-05). Total testosterone was correlated with obesity-related phenotypes in opposite directions in women (positive) and men (negative), including body fat percentage ($r_g$-female (F)=0.172 (0.0212), $r_g$-male (M)=-0.320 (0.0247)), body mass index ($r_g$-F=0.170 (0.0195), $r_g$-M=-0.302 (0.0207)), waist circumference (rG-F=0.163 (0.0205), $r_g$-M=-0.344 (0.0215)), and hip circumference (rG-F=0.148 (0.0217), rG-M=-0.263 (0.0226)). Instead, Testosterone-M was positively correlated with general well-being, as indicated by: not being on medication for cholesterol, blood pressure, diabetes, etc. ($r_g$=0.302 (0.0307)), recent strenuous physical activity ($r_g$=0.190 (0.0340)), and forced vital capacity ($r_g$=0.148 (0.0226)) (Supp. Figure 8).

#### Tissue- and cell-type enrichment of testosterone heritability

As GTEx datasets do not comprehensively represent female reproductive tissues of interest, particularly the ovary, we curated gene sets for ovarian cell types from publicly available single-cell gene expression databases[129,130]. At FDR<5%, we find enrichment of testosterone-F across a range of ovarian endothelial and immune cell types, as well as granulosa cell progenitors (*P*=0.0106), granulosa cumulus cells (*P*=0.0113), and theca cells (*P*=0.00176) (Supp. Figure 10).

#### Directional selection at testosterone-associated loci

We found evidence of recent directional selection, as measured by extreme SDS (in the lowest 0.25$^{th}$ %ile and highest 99.75$^{th}$ %ile), at four loci associated with testosterone (Supp. Figure 9). We observed negative selection of the testosterone-M-increasing allele at *DNAL1*, where we found that variants with higher probability of association with testosterone also had more negative selection scores. While we observed positive selection of the testosterone-increasing alleles at three loci: female-specific associations at the *CXCR4* and *SLC17A1* loci, and male-specific associations at the *SRD5A2* locus, the relationship between strength of variant association with testosterone and selection score was not consistent.

#### Reproductive hormone-associated genes implicated by gene burden analyses

Aside from the novel *HSD11B1* finding for testosterone-F, we report eight additional genes associated with testosterone-F at exome-wide significance (*P*<5E-06) that have not previously been implicated in GWASs, including those expressed in the ovaries (*ANAPC2*) and adrenal glands (*GPC2*), and genes associated with metabolism (*PDE3B*, *TAP2*, *NOTCH4*, and *ZKSCAN1*) (Supp. Figure 11 and Supp. Table 14). We also identified, for the first time, the *ELFN2* association with testosterone-M (Supp. Figure 11 and Supp. Table 14). Finally, across the reproductive hormones studied here, we replicated genes that have previously been reported to carry either rare or common variants associated with: FSH-F (genes *CHEK2* and *DCLRE1A*), Oestradiol-F (gene *SHBG*), Testosterone-F (18 genes, including two that are novel for testosterone but have known estrone associations), and Testosterone-M (22 genes, including eight that are novel for testosterone but have known sex-hormone binding globulin associations) (Supp. Figure 11 and Supp. Table 14).

##### Concordance between rare and common genetic architecture for testosterone

In the majority (83%) of genes with testosterone-associated rare variants, the rare variant has a larger absolute effect size on the trait than do any common variants in the gene. For example, a rare missense variant (chr17:7631360:C:T, MAF=0.612%) in *SHBG* is associated with large effects on testosterone-M (β=-0.743, *P*=1.69E-291), testosterone-F (β=-0.121, *P*=7.83E-09), and oestradiol-F (β=-0.259, *P*=1.03E-09); the common variants in *SHBG*, have effect sizes smaller by a factor of 7.5 (largest absolute effect of a common variant in *SHBG* on testosterone-M: β=-0.197, *P*=3.46E-82, testosterone-F: β=-0.0324, *P*=6.55E-22, and oestradiol-F: β=-0.035, *P*=2.67E-11) (Supp. Table 15).

##### Increased risk of infertility in individuals carrying rare testosterone-associated variants

In addition to the *GPC2* variant associated with lower testosterone and higher risk of infertility in women, we find a nominally significant association between a testosterone-lowering missense variant in *ZAN* and female infertility (chr7:100766559:A:G, F-ALL OR=1.37 (0.949-1.98), *P*=0.0463) (Figure 6B). *ZAN* is expressed in the testis and knockout mice display impaired sperm binding to the zona pellucida[149]. In men, a testosterone-lowering damaging missense variant in *HNF4A* (chr20:44413714:C:T OR=17.6 (2.26-137.07), *P*=3.08E-03) was associated with increased odds of infertility (Figure 6B).

#### Supplementary Discussion

Although infertility is a highly prevalent condition, the complex and heterogeneous causes of this condition remain poorly understood[1]. The case proportion of male and female infertility was between 0.3% and 1.4% in UKBB, which may reflect: (1) the biases of UKBB for healthy participants[150], (2) participants who were born between 1950-70 and thus reached reproductive age when infertility treatments were not widely available[151], and (3) the separation of fertility treatment records from other medical databases in the UK[151]. On the other hand, the Estonian and Danish data more closely reflect the population prevalence of infertility (between 7.0% to 13.2%) - the Estonian biobank is currently recruiting participants and is reflective of the current age distribution in Estonia[152], with recent infertility diagnoses included in the dataset, while the Copenhagen Hospital Biobank consists of patients with blood draws in Danish hospitals[153] and is thus more likely to capture individuals who have interacted with the healthcare system, which is genetically correlated with infertility in our study.

While we did not identify any genetic loci for infertility that have previously been reported for educational attainment or behavioural traits, we observed genetic correlations between female infertility and traits correlated with educational achievement[154,155], which is itself associated with delayed age at first birth[156]. There may still be residual confounding of age in our phenotype for female infertility of all causes.

Rare protein-coding variants captured in whole exome sequencing studies are valuable in revealing the underlying biology of phenotypes, as they directly implicate effector genes[157]. Indeed, in our UK Biobank-based WES analyses, we found that several enzymes involved in steroid hormone synthesis and metabolism, such as those in the hydroxysteroid dehydrogenase (HSD) family[82], carry rare variants that affect reproductive hormone levels. We also displayed concordance between the testosterone-altering effects of common and rare variation in a gene, mirroring the inverse relationship between allele frequency and effect size that has been observed across several other phenotypes[158].

Testosterone is among the most sexually dimorphic phenotypes in humans and has a corresponding dimorphic genetic architecture. Previous studies[159] and ours estimate the genetic correlation between female-specific and male-specific testosterone to be negligible (between 0 to 3%), highlighting the importance of disaggregating both genetic and phenotypic analyses by sex. We also found opposing associations of total testosterone with adiposity in the two sexes - testosterone in women is genetically correlated with higher overall and abdominal obesity, as measured by BMI, WC and HC, body fat percentage, fat mass, etc., but these were all negatively correlated with total testosterone in men. These sexually dimorphic genetic correlations reflect reported observational and causal associations in the literature[160,161], and may be related to the hormonal activity of adipose tissue, which metabolises steroid hormones and is differently distributed in men and women[162]. As testosterone and sex-hormone binding globulin are thought to partially mediate the deleterious cardiovascular consequences of increased visceral adipose tissue mass in men and post-menopausal women[163,164], understanding the shared genetic aetiology of testosterone and adiposity could lead to novel insights and treatments for cardiovascular disease.

Indeed, despite our best efforts to encompass heterogeneity in the causes and effects of infertility across the globe, we remained under-powered to perform sub-type specific GWASs for different categories of male infertility, where diagnosis and treatment lags behind female infertility[165]. Further, while the all-ancestry GWAS meta-analyses included one cohort of South Asian ancestry (Genes and Health) for infertility meta-analyses, and individuals of African, East Asian, and South Asian ancestries in the UKBB for hormone meta-analyses, over 90% of participants in our GWASs were of European ancestry. Even within the European-ancestry analyses, we demonstrated the importance of diverse population representation for biological insights in genetic studies. We identified six infertility-associated variants that were common in the Finnish and Estonian populations (MAF>1%), but not present in any of the other European cohorts, some of which may have immediate implications for fertility - such as the male infertility variant near *ENO4*, a gene expressed in the testis and involved in sperm motility[44,45]. Increasing the ancestral diversity in GWASs will not only be of great benefit to populations with ancestry-specific variants that affect fertility and reproductive hormone levels, but may also reveal shared novel biology.

### Supplementary Methods

#### Study populations for GWAS meta-analysis

**UK Biobank.** The UK Biobank (UKBB) is a prospective UK-based cohort study with approximately 500,000 participants aged 40–69 years at recruitment for whom a range of medical, environmental, and genetic information is collected[166]. Genotyping using two custom Affymetrix arrays, initial genotype quality control (QC), and imputation to the hg19 reference genome were performed by UKBB[167]. Sample ancestry was genetically ascertained as outlined below; in total, we retained 487,202 individuals of European ancestry for case-control analyses (Supp. Table 2), and 394,378 individuals of European ancestry, 10,548 individuals of African ancestry, 1,079 individuals of East Asian ancestry, and 7,019 individuals of South Asian ancestry with at least one hormone measurement for quantitative trait analyses (Supp. Table 9). GWASs in the UKBB were additionally adjusted for one-hot encoding of data provider or assessment centre, genotyping batch, genotyping array, and the first 21 genetic principal components (PCs).

We assigned sample population labels by training a random forest (RF) classifier using the 1000 Genomes 'super-population' labels. We first ran principal components analysis (PCA) on unrelated individuals in the 1000 Genomes project dataset, subset to LD-pruned autosomal variants. Samples in the UKBB genotyping data are projected onto this PCA space, ensuring that we correctly account for shrinkage bias in the projection[168,169]. Next, we used the 'super-population' labels (AFR=Africans, AMR=Admixed Americans, EAS=East Asians, Europeans=EUR, South Asians=SAS) of the 1000 Genomes dataset to train a RF classifier, using the randomForest (4.6) library in R[170], and predicted the super-population for each of the UKBB samples. Samples with classification probability >0.99 were retained for downstream analysis.

**Avon Longitudinal Study of Parents and Children.** The Avon Longitudinal Study of Parents and Children (ALSPAC) is a longitudinal population-based study that recruited 13,761 pregnant women resident in the South West of England, with expected delivery dates between 1 April 1991 and 31 December 1992, who have continued to be followed up over the last 32 years[171]. GWASs were performed on reproductive hormone values measured in follow-up assessments (not during pregnancy) from approximately 3,000 mothers with linked genetic data. Genome-wide genotyping was performed using the Illumina HumanHap550 quad chip genotyping platform at 477,482 markers and imputed to the hg19 reference genome using the Haplotype Reference Consortium (HRC) panel[171,172]. GWASs in ALSPAC were performed using the BOLT-LMM software[173] and additionally adjusted for the first 10 genetic PCs.

**Copenhagen Hospital Biobank & The Danish Blood Donor Study.** The Copenhagen Biobank (CHB) includes samples drawn for blood type testing or antibody screening from more than 500,000 hospitalised patients and outpatients in Danish hospitals in the Capital Regions[153]. The Danish Blood Donor Study is a prospective cohort study of 163,000 healthy blood donors. Genotyping in both cohorts was performed by deCODE Genetics using the Illumina Global Screening Array (GSA) microchip at 660,000 markers and imputed to the hg38 reference genome using a custom panel[153,174]. The GWASs in CHB/DBDS were performed with SAIGE v1.18 were additionally adjusted for the first 10 genetic PCs.

**deCODE.** 173,025 Icelanders that have been genotyped using Illumina SNP chips were long-range phased[175] and variants identified in the WGS of 63,460 Icelanders were imputation into chip-typed individuals and their close non-chip-typed family members[176]. The sequencing was done using Illumina standard TruSeq methodology. Only samples with a genome-wide average coverage of 20X were considered. Autosomal SNPs and INDEL's were called using Graphtyper version 1.4.5. Variants that did not pass quality control were excluded from the analysis. Information about haplotype sharing was used to improve variant genotyping, taking advantage of the fact that all sequenced individuals had also been chip-typed and long-range phased.

Information on the fertility phenotypes was extracted from information from phenotype records from questionnaires and healthcare centres. Logistic regression analysis assuming a multiplicative model was used to test for association[176] between fertility cases/control phenotypes, additionally adjusted for year of birth and county of origin. For hormone biomarkers, values were additionally adjusted for measurement centre and county of origin, standardised using an inverse-normal transform and tested for association with sequence variants using a generalised linear model. Any inflation in the test statistic due to population structure and relatedness of the participants was adjusted for using LD score regression.

**Estonian Biobank.** The Estonian Biobank (EstBB) is a population-based biobank of Estonia, reflecting the age, sex, and geographic distribution of the population, with a cohort size of 200,000 individuals (approximately 20% of the nation) with EHR linkage[152]. Individuals were genotyped using the Illumina Global Screening Array (GSA) microchip at 700,000 markers and imputed to the hg19 reference genome using a custom panel[177]. GWASs in EstBB were performed using the REGENIE software v3.0.3[107] and additionally adjusted for the first 10 genetic PCs.

**FinnGen.** The FinnGen study is a large-scale genomics initiative that has analyzed over 500,000 Finnish biobank samples and correlated genetic variation with health data to understand disease mechanisms and predispositions. The project is a collaboration between research organisations and biobanks within Finland and international industry partners. FinnGen Data Release R10 was used in the present study. Individuals were genotyped with Illumina and Affymetrix chip arrays, and genotype data were imputed using the population-specific SISu v4.2 imputation reference panel of 8,554 whole genomes. GWAS was performed using the REGENIE software v2.2.4[107] and additionally adjusted for the first 10 genetic PCs and genotyping batch.

**Genes and Health.** The Genes and Health study (G&H) is a longitudinal prospective cohort of participants of British Bangladeshi and Pakistani origin recruited from East London, Bradford, and Manchester, with EHR linkage[178]. Participants were genotyped using the Illumina GSA Multi-Disease GSAv3EAMD chip and imputed to the hg38 reference genome using the TOPMED reference panel[178,179]. GWASs in G&H were performed using the REGENIE software[107] and additionally adjusted for the first 20 genetic PCs.

**Publicly available summary statistics for reproductive hormones.** We downloaded publicly available GWAS summary statistics from studies without UK Biobank participants for FSH and LH[180,181,182], progesterone[183], oestradiol[183], and testosterone[181,183,184] from the GWAS Catalog[62] (Supp. Table 9). Summary statistics from Suhre *et al.* (2017)[180] were missing non-effect alleles, which were merged in by matching chromosome and genome position information from Ensembl hg19 variant call files[134]. These and the summary statistics from Prins *et al.* (2017)[184] were missing allele frequencies, which were merged in from chromosome, position, and allele-matched frequencies derived from European-ancestry individuals in the 1000 Genomes dataset[112]. All summary statistics on genome build hg19 were lifted over to hg38 using the UCSC liftOver tool[185] - between 97.3% and 99.7% of variants were successfully lifted over to the hg38 genome assembly.

##### Exome sequencing variant and sample-level QC

We defined "high quality" variants as those MAF>0.001 and call rate ≥0.99 falling within the UK Biobank capture intervals plus 50 bp padding. These variants were used to evaluate sample-level metrics of mean call rate and depth. We retained samples satisfying all of the following:

- Genetic sex inferred as XX or XY
- Mean call rate ≥0.99.
- Mean coverage ≥20x.
- Not withdrawn.

Next, we removed variants satisfying at least one of the following criteria:

- The variant lies outside the UK Biobank capture plus 50 bp padding.
- The variant lies within a low complexity region.
- The variant lies within a segmental duplication.

Among this (sample, variant) set, we ran Hail's `sample_qc()`[186] to remove samples lying outside the median ± 4 median absolute deviations (MADs) within each super-population (see "Assigning population labels" in Methods section above). The QC protocol was split by UK Biobank WES tranche (50k, 200k, 450k) to guard against batch effects, as tranches were sequenced in separate runs. The following metrics were used for QC:

- Number of deletions (`n_deletion`).
- Number of insertions (`n_insertion`).
- Number of SNPs (`n_snp`).
- Ratio of insertions to deletions (`r_insertion_deletion`).
- Ratio of transitions to transversions (`r_ti_tv`).
- Ratio of heterozygous variants to homozygous alternate variants (`r_het_hom_var`).

Following MAD filtering (Supp. Figure 12, Supp. Table 18), 402,345 European samples were retained for analysis. For each sample, we excluded non-passing sites as described in Karzcewski *et al.* (2022) [135]. Briefly, an RF classifier was trained to distinguish true positives from false positive variants using a collection of allele and site annotations. Variants were assigned 'PASS' to maximise sensitivity and specificity across a series of readouts in trio data and precision-recall in two truth samples, after which samples with excess heterozygosity (defined as inbreeding coefficient<−0.3) were removed. Next, we removed low quality genotypes by filtering to the subset of genotypes with depth≥10 (5 among haploid calls), genotype quality≥20, and minor allele balance >0.2 for all alternate alleles for heterozygous genotypes. Following this filter, we remove variants that were not called as "high quality" among any sample.
